# Bidirectional contact tracing dramatically improves COVID-19 control

**DOI:** 10.1101/2020.05.06.20093369

**Authors:** William J. Bradshaw, Ethan C. Alley, Jonathan H. Huggins, Alun L. Lloyd, Kevin M. Esvelt

## Abstract

Contact tracing is critical to controlling COVID-19, but most protocols only “forward-trace” to notify people who were recently exposed. Using a stochastic branching-process model, we show that “bidirectional” tracing to identify infector individuals and their other infectees robustly improves outbreak control, reducing the effective reproduction number (*R*_eff_) by at least ∼0.3 while dramatically increasing resilience to low case ascertainment and test sensitivity. Adding smartphone-based exposure notification can further reduce *R*_eff_ by 0.25, but only if nearly all smartphones can detect exposure events. Our results suggest that with or without digital approaches, implementing bidirectional tracing will enable health agencies to control COVID-19 more effectively without requiring high-cost interventions.

## Introduction

Contact tracing, isolation, and testing are some of the most powerful public health interventions available. The nations that have most effectively controlled the ongoing COVID-19 pandemic are noteworthy for conducting comprehensive and sophisticated tracing and testing^1^. Current “forward-tracing” protocols seek to identify and isolate individuals who may have been infected by the known case, preventing continued transmission (Fig. 1a). For example, the European Union and World Health Organization call for the identification of contacts starting two days prior to the development of symptoms^2,3^.

**Figure 1.**
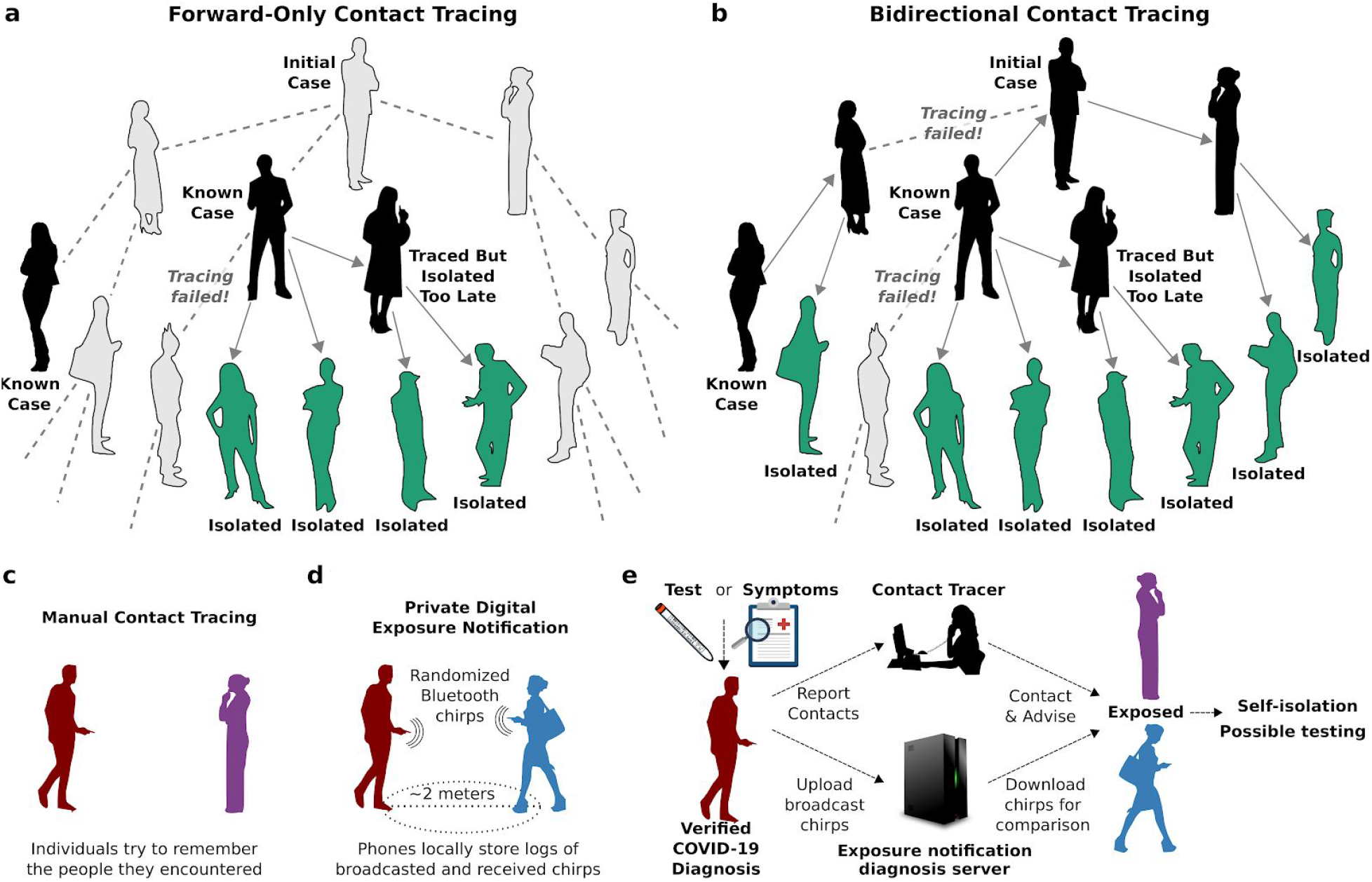
Forward-only and bidirectional contact tracing and digital exposure notification. (a) Notifying people exposed to known cases (black) and isolating them (green) can prevent further transmission, but will miss asymptomatic and undiagnosed cases (gray) and descendants. (b) Bidirectional tracing also notifies and tests potential infectors, enabling isolation of additional cases. (c) Manual contact tracing requires individuals to share recent contacts with health authorities. (d) In digital exposure notification, smartphones broadcast rotating pseudorandom “chirps” and record those emitted by nearby devices^11^. (e) Individuals diagnosed with COVID-19 can “opt-in” by uploading broadcasted chirps to a diagnosis server^11^. All devices frequently check the server and alert the user if calculated exposure exceeds a threshold set by the local health authority. In hybrid manual+digital systems, human tracers would seek to identify contacts without smartphones.

However, chains of SARS-CoV-2 transmission may persist despite excellent medical monitoring and forward-tracing programs due to substantial rates of undiagnosed or asymptomatic transmission^4^ (Fig. 1a). Asymptomatic carriers, who reportedly bear equivalent viral loads to patients exhibiting symptoms^5^, have been estimated to account for 18%^6^ to 79%^7^ of cases, with multiple population surveys indicating intermediate values around 45%^8–10^.

We hypothesized that when asymptomatic carriers are common, “bidirectional” contact tracing could identify and isolate undiscovered branches of the transmission tree, preventing many additional cases (Fig. 1b). Bidirectional contact tracing uses “reverse-tracing” to identify the parent case who infected a known case, then continues tracing to iteratively discover other cases related to the parent. It has been successfully used to identify clusters and community transmission in Japan^12^ and Singapore^13,14^, but is otherwise uncommon. Because the incubation time for SARS-CoV-2 is longer than 48 hours, existing tracing protocols cannot identify parent cases^2,3^, and previous studies of COVID-19 contact tracing have neglected the possibility of gains from bidirectional tracing.

Numerous ongoing efforts aim to use smartphones emitting randomized Bluetooth and/or ultrasound “chirps” to notify people exposed to infected individuals (Fig. 1d–e). Digital approaches may offer considerable advantages in speed^15^, scale, efficacy^4^, and confidentiality^16^. To investigate the efficacy of bidirectional tracing, we adapted and extended a stochastic branching-process model of SARS-CoV-2 forward-tracing^17^ and used it to explore the efficacy of different tracing strategies under plausible epidemiological scenarios.

## Results

### Bidirectional tracing improves epidemic control in manual and idealized digital scenarios

In our model, each case generates a number of new cases drawn from a negative binomial distribution, with incubation and generation-time distributions based on the published literature (Table 1 & Table S1). Cases could be identified and isolated based on symptoms alone or through contact tracing (Methods). Each outbreak was initialized with 20 index cases to minimize stochastic extinction, and designated as “controlled” if it reached extinction (zero new cases) before reaching 10,000 cumulative cases.

**Table 1:** Key parameters of the branching-process model.

| Parameter | Value |  |  | Sources and Notes |
| --- | --- | --- | --- | --- |
|  | Median | Optimistic | Pessimistic |  |
| % asymptomatic carriers | 45% | 40% | 55% | 8–10,37 |
| Relative infectiousness of asymptomatic carriers | 50% | 45% | 60% | Informed by viral loads and tracing results described in <sup>5,9,26,30–32</sup> |
| % environmental transmission | 10% | 5% | 15% | <sup>4,38</sup> |
| Proportion of pre-symptomatic transmission | 48% | 38% | 53% | Informed by <sup>5,26,32,39–44</sup> |
| % of cases with chirping smartphones | 53% (low-uptake) / 80% (high-uptake) |  |  | Survey data <sup>22,23,45</sup> |
| Test sensitivity | 80% |  |  | <sup>20,21</sup> |
| $R_0$ (default) | 2.5 | | | Most estimates cluster between 2.0 and 3.0: <sup>4,9,25–28</sup> |
| Incubation period | 5.5 ± 2.1 days (lognormal distribution) |  |  | <sup>46</sup> |
| Delay from onset to isolation | 3.8 ± 2.4 days (Weibull distribution) |  |  | <sup>19</sup> |
| Delay for testing | 1 ± 0.3 days (gamma distribution) |  |  | Assumed |
| Delay for manual tracing | 1.5 ± 4.8 days (lognormal distribution); median 0.5 days |  |  | Previous reports suggest most contacts can be traced within one day, but some take much longer <sup>47</sup> |
| Delay for digital tracing | 0 days |  |  | Assumed |

We began by investigating a median scenario in which 10% of transmission was assumed to be environmental (and therefore untraceable), 48% of transmission occurred pre-symptomatically, and 45% of cases were asymptomatic with reduced (50%) infectiousness. For the initial analysis we assumed a fixed basic reproduction number (*R*_0_) of 2.5 (Fig. 2), but explored other *R*_0_ values below.

**Figure 2.**
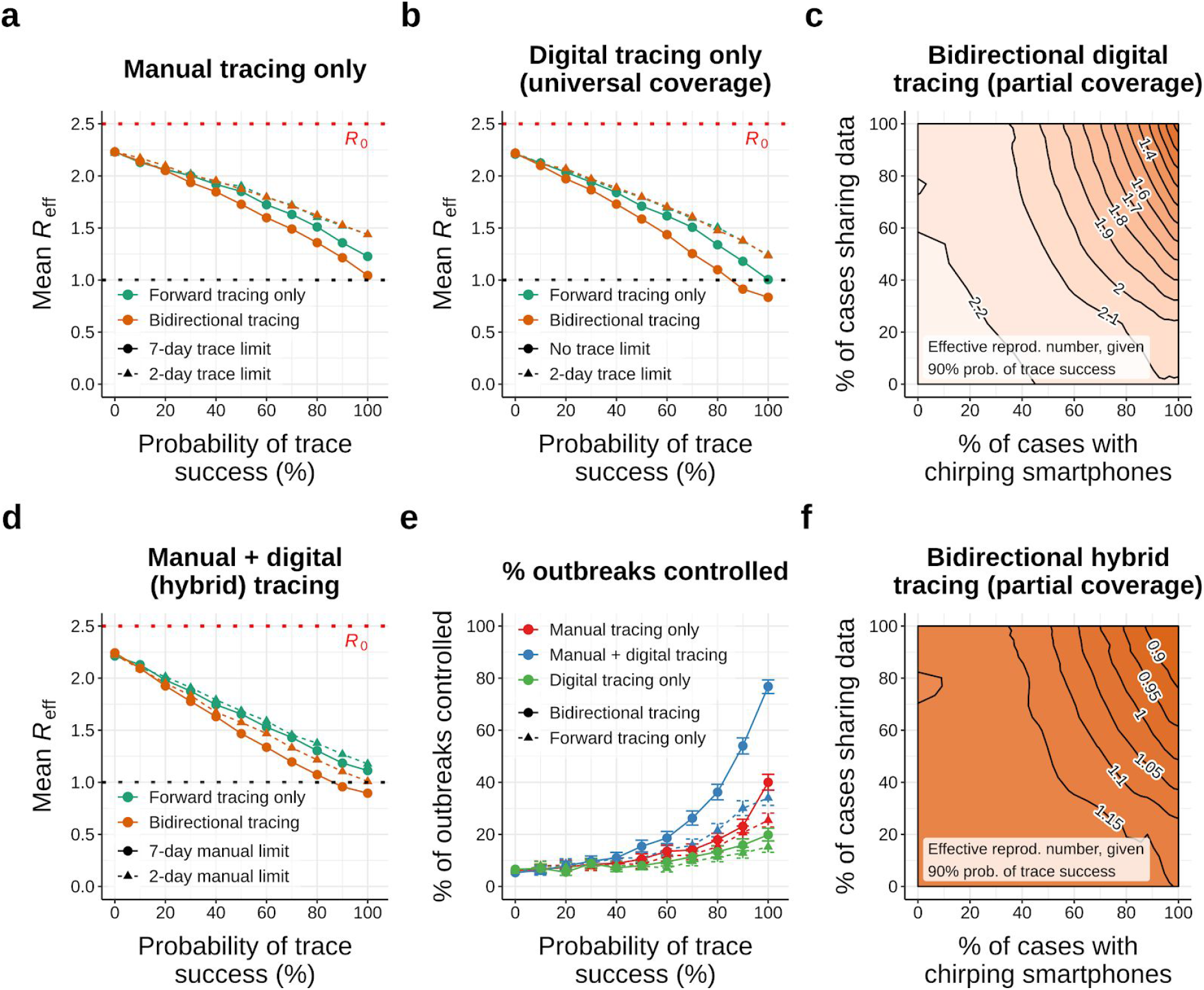
Comparing forward and bidirectional contact tracing at *R*_0_ = 2.5. (a) Average R_eff_ achieved by manual tracing with 2-day and 7-day manual tracing windows. (b) Average R_eff_ achieved using digital tracing alone, assuming universal smartphone coverage and data sharing. (c) Neighbor-averaged contour plot depicting R_eff_ for digital-only tracing with varying smartphone coverage and data-sharing, assuming 90% probability of trace success. (d) Average R_eff_ achieved using hybrid manual+digital tracing, assuming 90% data sharing and 80% smartphone usage. (e) Percentage of outbreaks controlled under manual, digital and hybrid tracing, assuming 90% data sharing, 80% smartphone usage, and a 7-day manual tracing window. (f) Neighbor-averaged contour plot of R_eff_ for hybrid tracing, assuming a 7-day manual tracing window and 90% probability of trace success. All panels assume median disease parameters (Table S1). “Probability of trace success” refers to trace attempts that are not otherwise blocked by environmental transmission or fragmentation of the digital network. Points in (a,b,d) and contours in (c,f) represent average values across 1000 runs; error bars in (e) represent 95% credible intervals under a uniform prior.

In this scenario, manual forward tracing of contacts occurring up to 48 hours before symptom onset per current guidelines^2^ reduced *R*_eff_ as anticipated, but not nearly enough to control the pandemic without other measures: even if all non-environmental contacts within that time window were successfully traced, *R*_eff_ remained in excess of 1.4 (Fig. 2a). Extending the tracing window to 7 days before symptom onset yielded a moderate improvement, reducing the best-case *R*_eff_ by roughly 0.2 (Fig. 2a and Fig. S1); further widening of the tracing window resulted in minimal additional benefit (Fig. S2). Switching from forward-only to bidirectional tracing offered a further gain of similar magnitude, bringing the best-case *R*_eff_ close to the critical threshold of 1.0. Even here, however, the fraction of outbreaks controlled was less than 50% (Fig. 2e).

Compared to manual tracing, digital exposure notification is both faster and scalable to much wider time windows. When all contacts in the past 14 days were available for analysis^11^, idealized digital forward-tracing produced markedly superior outcomes relative to manual forward-tracing (Fig. 2b), in agreement with earlier models^4^. Crucially, bidirectional digital tracing exhibited an even more dramatic improvement over the forward-only approach, successfully bringing *R*_eff_ below 1.0 without any other control measures (Fig. 2b) and more than doubling the best-case probability of control (Fig. S5).

### Digital tracing is fragile to network fragmentation

Idealized digital contact tracing appears promising, but assumes all individuals carry exposure-detecting smartphones and upload their broadcasted chirps when diagnosed with COVID-19. We hypothesized that the effectiveness of digital tracing would rapidly degrade when fewer people participated, in line with or worse than the quadratic dependence noted by others^4,16,18^.

As predicted, even small decreases in the proportion of individuals carrying a participating smartphone or (to a lesser extent) sharing their broadcasted chirps resulted in fragmentation of the tracing network (Fig. 2c, and Fig. S3 & S6), increasing *R*_eff_ to levels comparable to, or even worse than, manual tracing alone. As a consequence of this fragility, our results suggest that digital tracing alone cannot currently substitute for traditional manual tracing, even under very optimistic assumptions about uptake and use^19,20^.

### Hybridizing manual and digital tracing improves performance

Neither manual nor digital contact tracing alone sufficed to control COVID-19 in our median scenario. Digital tracing is fast and comprehensive but highly fragile to network fragmentation; manual tracing is slower and limited to a narrow time window, but more robust. We hypothesized that the two methods could complement each other, with manual contact tracers focusing their effort on tracing contacts invisible to the digital system, and that this hybrid approach might outperform either method in isolation.

When 80% of cases participated in the digital system, supplementing bidirectional manual with digital tracing substantially improved epidemic control (Fig. 2d), reducing best-case *R*_eff_ by roughly 0.15 compared to manual tracing and more than 0.5 compared to digital tracing with equivalent uptake. Compared to manual tracing, a hybrid approach roughly doubled the probability of controlling individual outbreaks (Fig. 2e). Control under hybrid tracing was still somewhat sensitive to uptake of the digital system (Fig. 2f); but far less so than digital tracing alone. Performing only forward-tracing, or reducing the width of the manual tracing window, substantially degraded performance (Fig. 2d–e & Figs. S3–S7).

### Bidirectional tracing is robust to low case ascertainment and test sensitivity

Our results thus far assume that 90% of symptomatic cases can be identified based on symptoms alone, corresponding to an overall ascertainment rate (including asymptomatic cases) of roughly 50%. Unsurprisingly, reducing the percentage of symptomatic cases so identified impaired epidemic control (Fig. 3a & Fig. S31). However, bidirectional tracing was far more robust to these changes than forward-only tracing, resulting in dramatically lower R_eff_ values across a wide range of ascertainment rates.

**Figure 3.**
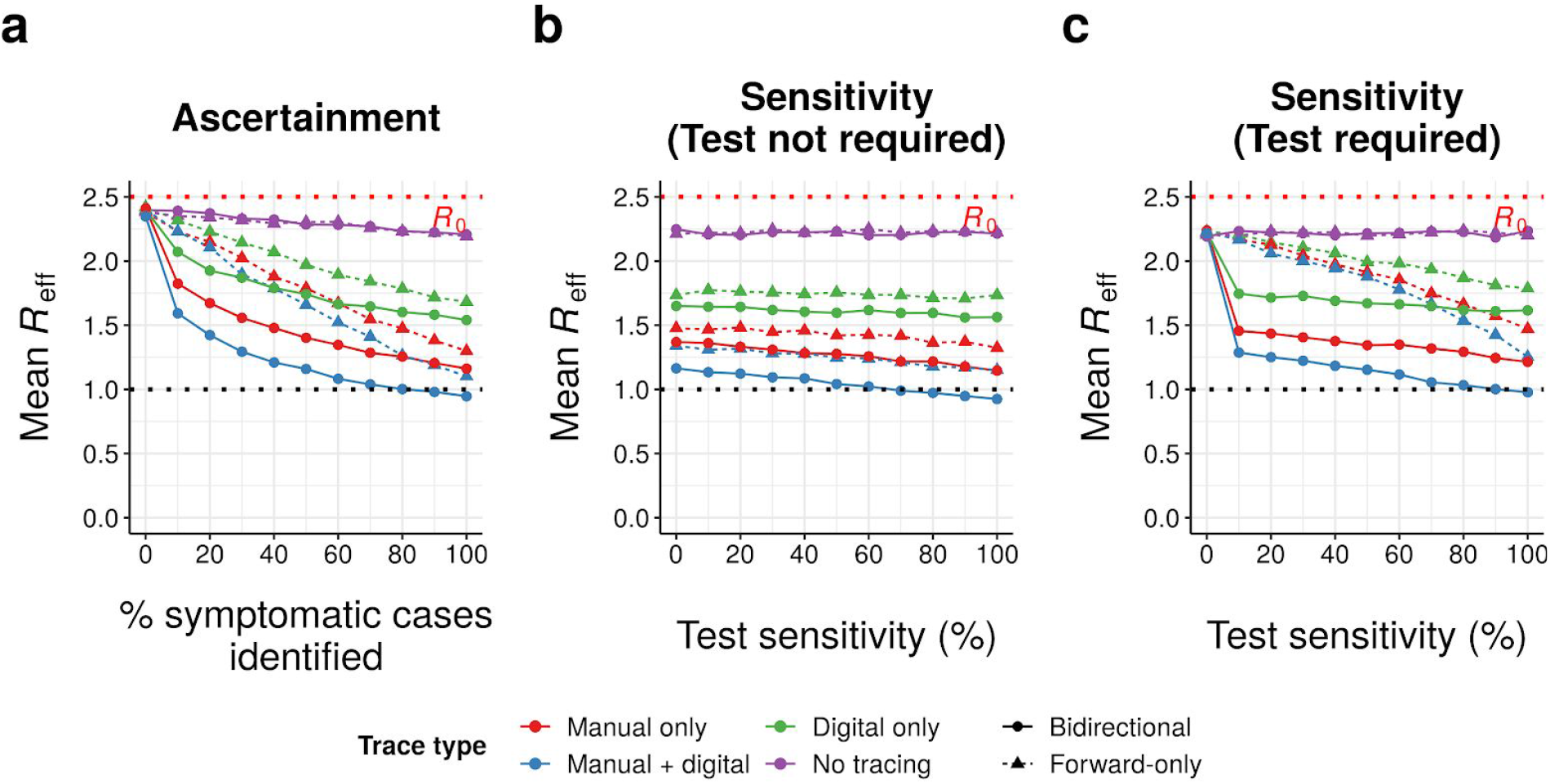
Bidirectional tracing under reduced ascertainment and test sensitivity. Mean effective reproduction number (*R*_eff_) achieved across 1000 runs per scenario by different tracing strategies (manual, digital, both or neither; forward-only or bidirectional) as a function of (a) the percentage of symptomatic cases that can be identified by health authorities based on symptoms or (b-c) test sensitivity, assuming 90% probability of trace success, a 7-day manual tracing window, high-uptake digital tracing, and median disease parameters (Table S1). In (a-b), tracing can be initiated from symptomatic cases without a test, while in (c) a positive test result is required; in both cases, a positive test result is required to initiate tracing from pre- or asymptomatic cases. (a) assumes test sensitivity of 80%, while (b-c) assume 90% of symptomatic cases are identified on the basis of symptoms. “Probability of trace success” refers to trace attempts that are not otherwise blocked by environmental transmission or fragmentation of the digital network.

When symptomatic cases can be traced immediately based on symptoms alone (our default assumption),, both forward-only and bidirectional tracing were fairly robust to deviations in test sensitivity from our default assumption of 80%, with *R*_eff_ values increasing only slowly as sensitivity falls (Fig. 3b). In contrast, if a positive test result was required before tracing from symptomatic cases (as is the case in many countries^21^), the efficacy of forward-tracing became dramatically more dependent on a high test sensitivity: low sensitivities yielded greatly increased R_eff_ values (Fig. 3c & Fig. S30), consistent with previous modeling studies reporting impaired performance under these conditions^21^. In sharp contrast, the performance of bidirectional tracing remained relatively robust to changes in test sensitivity even under these conditions.

### High-uptake hybrid bidirectional tracing robustly doubles the probability of outbreak control

To evaluate the epidemiological robustness of our findings, we repeated our analysis using *R*_0_ values ranging from 1.0 to 4.0 (Fig. 4, left)^4,9,22–24^. We assumed tracing of 90% of non-environmental contacts, a 7-day manual trace window, immediate tracing of symptomatic cases, and high uptake of the digital system. A wider range of assumptions are explored in Figures S9–S31.

**Figure 4.**
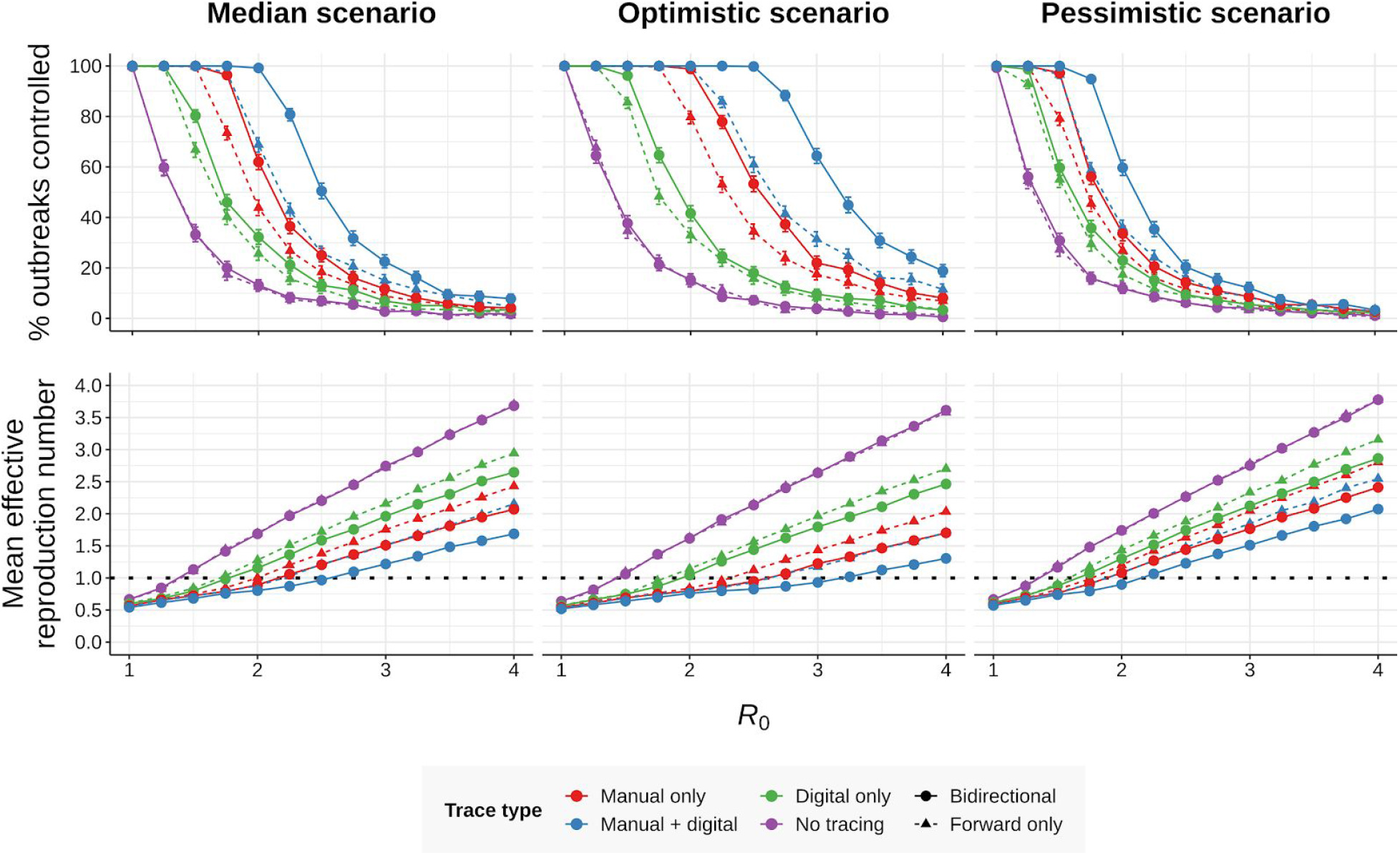
Effect of *R*_0_ and disease parameters on performance. (top row) Percentage of outbreaks controlled and (bottom row) average effective reproduction number under various forms of contact tracing (manual, digital, both or neither; forward-only or bidirectional) as a function of the basic reproduction number *R*_0_, assuming (left) median, (middle) optimistic or (right) pessimistic disease parameters (Table S1), 90% non-environmental contacts traced, a 7-day manual trace window, and high-uptake digital tracing. Error bars in the top row represent 95% credible intervals across 1000 runs under a uniform beta prior; points in the bottom row represent average values over the same. Isolating symptomatic cases can dampen the outbreak at low *R*_0_, even in the absence of tracing.

Small reductions in *R*_0_ resulted in large increases in control probability across all forms of tracing (Fig. 4, left). Hybrid bidirectional tracing robustly outperformed all other tracing strategies, with the greatest degree of outperformance observed when 1.75 ≤ *R*_0_ ≤ 3.5. When *R*_0_ ≤ 1.75, manual and hybrid tracing both achieved nearly 100% control, while when *R*_0_ ≥ 3.5, no strategy achieved control probabilities over 10%. Even in these low-control scenarios, however, hybrid bidirectional tracing consistently reduced R_eff_ by roughly 20% relative to manual tracing alone (Fig. 4 & Figs. S19–S20, S22, S32). Constraining manual tracing to a 2-rather than 7-day window substantially impaired performance (Fig. S10), while lowering uptake of the digital system from 80% to 53% of cases – in line with existing survey data^19–21^ and current plans for opt-in participation^11^ – mostly abrogated the advantage of hybrid over manual approaches (Figs. S16 and S21–S22).

While COVID-19 is clearly a challenging disease to control, there remains substantial uncertainty around the exact rates of asymptomatic, presymptomatic, and environmental transmission. To explore a wider range of scenarios, we aggregated our collective best estimates to define optimistic and pessimistic values for these parameters, with 5/15% environmental transmission, 38/53% pre-symptomatic transmission, and 40/55% asymptomatic carriers which were 45/60% as infectious as symptomatic cases. We repeated our simulations under these new assumptions for a range of *R*_0_ values (Fig. 4, middle & right).

While hybrid bidirectional tracing continued to robustly outperform other configurations (Figs. S17-S22), the probability of control varied substantially between scenarios: in the optimistic scenario, high-uptake hybrid bidirectional tracing was sufficient to reliably control outbreaks whenever *R*_0_ ≤ 2.5, while in the pessimistic scenario reliable control was only achieved at *R*_0_ ≤ 1.75. Restricting uptake of the digital system or the width of the manual tracing window impaired performance across all scenarios (Figs. S10 & S16-S22).

To summarize the predicted effects of different approaches, we compared the percentage of outbreaks controlled and *R*_eff_ values achieved under all three scenarios, in the absence of any other interventions (Fig. 5, & Figs. S33–S36). Relative to optimal current practice (i.e. 2-day forward-only manual tracing), 7-day bidirectional manual tracing achieved a reduction in *R*_eff_ of roughly 0.3 across all scenarios. Supplementing manual tracing with a low-uptake digital system provided a further reduction of 0.1 in the median and pessimistic scenarios, increasing to 0.25 (i.e., 0.55 total) when uptake was high. In total, switching from current practice to high-uptake hybrid bidirectional tracing approximately tripled the probability of controlling the spread of SARS-CoV-2 across all three scenarios.

**Figure 5.**
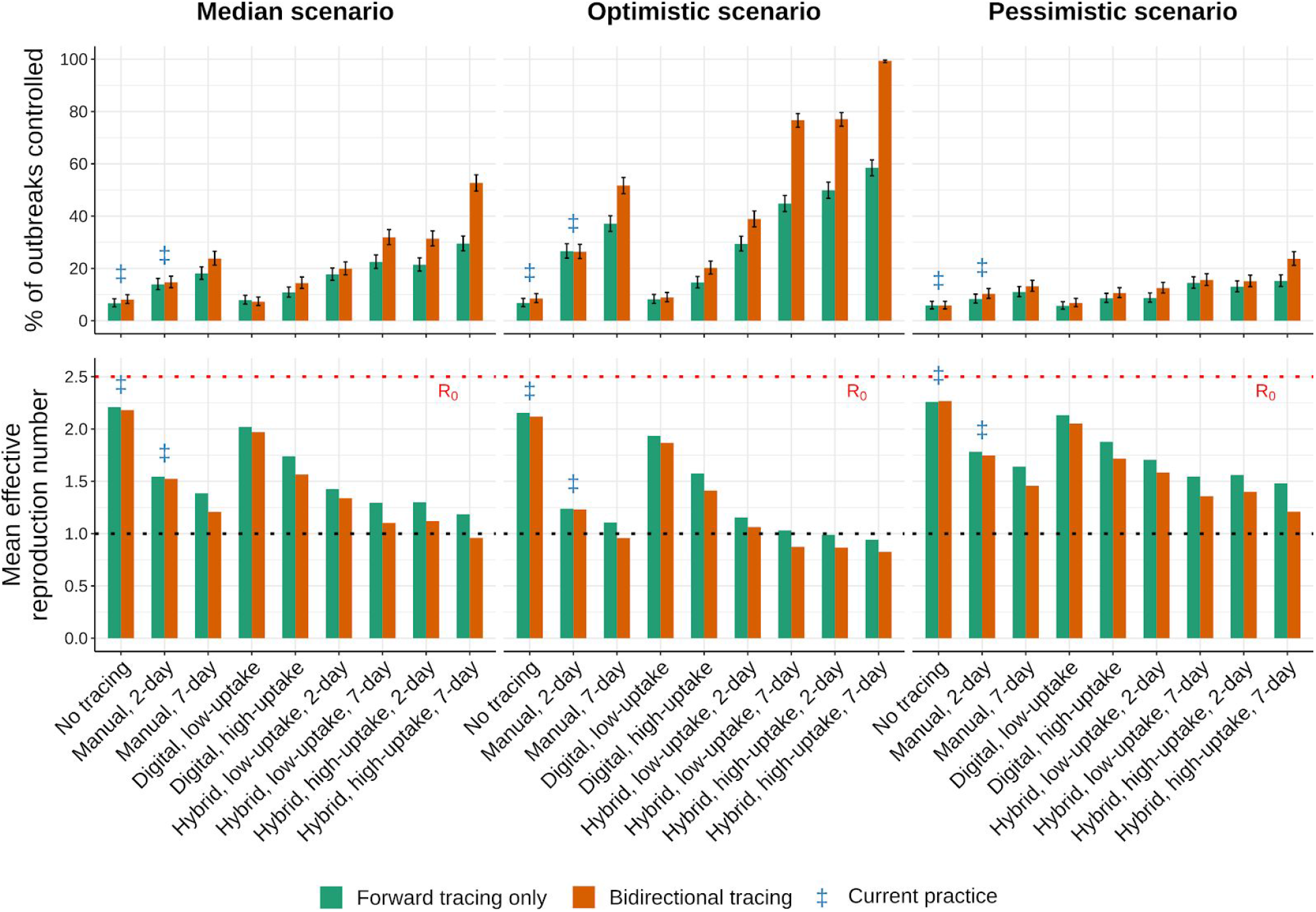
Performance of different tracing strategies relative to current practice. (top row) Percentage of outbreaks controlled, and (bottom row) mean effective reproduction number obtained under (left) median, (middle) optimistic, and (right) pessimistic scenarios (Table S1), assuming an *R*_0_ of 2.5, 90% of non-environmental contacts traced, 2- or 7-day manual trace windows, and immediate (pre-test) tracing of symptomatic cases. Blue ‡ symbols indicate current practice in most regions. Low and high uptake correspond to 53% and 80% of cases, respectively, having chirp-enabled smartphones. Error bars in the top row represent 95% credible intervals across 1000 runs under a uniform beta prior. Without tracing, forward and bidirectional are equivalent.

## Discussion

Given the tremendous suffering inflicted by the COVID-19 pandemic and the potentially critical role of expanding contact-tracing systems in its control, there is an urgent need to optimize the implementation of these systems.

Our model predicts that making tracing bidirectional would markedly improve COVID-19 control. Indeed, for a contact-tracing system to reach the levels of control cited by earlier studies^4,21^, we find that bidirectional tracing is *required*. Bidirectional tracing outperforms forward-tracing regardless of how the tracing is done. Simply switching to manual bidirectional tracing is sufficient to reduce *R*_eff_ by 0.3 if the time window for tracing is sufficiently wide, while high-uptake bidirectional hybrid tracing is predicted to be approximately three times as effective at controlling outbreaks as current best practice. The case for bidirectional tracing becomes even stronger when case ascertainment is otherwise suboptimal, or if a positive test result is required before tracing the contacts of a symptomatic case.

We stress-tested these conclusions with a wide range of plausible parameter combinations and possible values of *R*_0_. Notably, our “optimistic” scenario is more pessimistic than some earlier studies due to recent studies reporting higher values for both the rate and relative infectiousness of asymptomatic carriers^5,9,23,25,26^. Whether hybrid bidirectional tracing alone was sufficient to reliably control the pandemic was dependent on epidemiological parameters; however, if other low-cost precautions^27^ could reduce *R* _eff_ below 1.75, our model predicts that this strategy would bring transmission under control with high probability even under our pessimistic scenario.

Despite this stress-testing, our conclusions must be considered in the context of our model, which, while less idealized than its predecessors, has limitations. It makes no distinction between mild and severe symptoms, and does not consider demographic, geospatial or behavioural variation between cases. Since only true cases are included in the model, only the sensitivity of testing is considered; in reality, the balance between test sensitivity and specificity is a crucial trade-off, and high rates of false positives will severely impede response effectiveness and the credibility of the tracing system.

These limitations aside, there is considerable evidence that bidirectional tracing can be feasibly implemented in practice. Locales such as Singapore^13,14^ and Washington State^28^ have employed bidirectional tracing to determine whether community transmission is occurring, while Japan’s protocol explicitly aims to identify sources of infection^12^. The primary practical difference between contact with an infector and an infectee is the time at which the contact occurred; as such, the core obstacle to implementation in other areas is the cost of expanding the tracing window. Health authorities could accomplish this by expanding the workforce of contact tracers, leveraging the lightened workload afforded by a high-uptake digital system, or focusing limited resources on clusters as is done in Japan. Digital systems, which already track exposures for 14 days, can trace bidirectionally at no additional cost.

In addition to effective implementation of bidirectional tracing, a successful control program will also depend on the availability of timely COVID-19 testing^15^, high adherence to quarantine requests^4,17,29,30^, and scaling of manual contact tracing, while digital systems will require efficient algorithms with acceptable sensitivity and specificity. These caveats notwithstanding, our results indicate that bidirectional contact tracing could play an essential and potentially decisive role in controlling COVID-19 and preventing future pandemics.

## Data Availability

Code for configuring and running the model is publicly available at https://github.com/willbradshaw/covid-bidirectional-tracing.

## Acknowledgments

We thank Aaron Bucher of the COVID-19 HPC Consortium and Amazon Web Services for granting us extra cloud compute credits.

## Author Contributions

K.M.E. conceived the study. J.H.H. and A.L.L. identified a suitable model framework. W.J.B. designed and programmed the adapted model, advised by the other authors. All authors independently estimated key model parameters based on the literature. W.J.B. ran all simulations and generated figures with assistance from E.C.A. All authors jointly wrote and edited the manuscript.

## Methods

### Structure of the model - Infection dynamics

A new case is infected at some **exposure time**, equal to zero if the case is an index case and otherwise drawn from the **generation time distribution** of its parent case (see below). If not asymptomatic, the case develops symptoms at some **onset time** drawn from an **incubation time distribution**. Asymptomatic cases do not develop symptoms, but are still assigned an onset time for the purpose of determining their generation-time distribution (see below).

The number of child cases infected by the case is drawn from a negative binomial distribution, with mean equal to the appropriate reproduction number (see below) and heterogeneity determined by the overdispersion parameter *k*. The exposure times of these child cases are drawn from a skewed-normal **generation time distribution** centered on the symptom onset of their parent ^17^, with a skew parameter chosen to give a pre-specified probability of pre-symptomatic transmission (for a symptomatic parent) and an SD parameter of 2. The generation time distribution for an asymptomatic parent is centered on its “effective” onset time (see above). The shape of the generation-time distribution is the same for all cases.

The average number of children produced by a case depends on its symptomatic status, and is determined by the overall *R*_0_ value, the proportion of asymptomatic carriers *p*_*asym*_, and the relative infectiousness *x*_*asym*_of asymptomatic carriers (expressed as a fraction of *R*_0_). Given a reproduction number for asymptomatics of *R*_*asym*_ = *R*_0_ · *x*_*asym*_, the reproduction number of symptomatic cases that produces the desired overall *R*_0_is given by 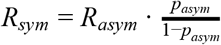

### Structure of the model - Infection control

Once symptoms develop, a case is **identified** by public health authorities with probability *p*_*isol*_, with the delay from onset to identification drawn from a **delay distribution**. Identified cases are instructed to isolate, and each case complies with that order with probability *p*_*comply*_. Cases that comply with isolation generate no further child cases after their time of identification. Asymptomatic cases cannot be identified from symptoms, but may be identified via contact tracing from other cases (see below); once identified, they are instructed to isolate as above. Tracing can also cause symptomatic cases to be isolated earlier than they would be from symptoms alone.

An identified case is **tested**, which takes time drawn from a test time distribution and returns a positive result with probability equal to the sensitivity of the test (since the model does not consider uninfected individuals, the specificity of the test is also not considered). For asymptomatic cases, or symptomatic cases identified prior to symptom onset, a positive test result is required to initiate contact tracing; symptomatic cases that have already developed symptoms can either be traced immediately upon identification, or require a positive test result prior to tracing, depending on model settings.

Whether before or after a test result is obtained, the contacts of an identified case can also be **traced**. Tracing can only proceed outward from a case if they share their contact history, either via a contact-tracing app or with a manual contact tracer (see below). Tracing can identify the children of the traced case (forward tracing), and may also be able to identify its parent (reverse/backward tracing), depending on model settings. The speed and success probability of tracing depend upon whether tracing is conducted digitally or manually, which in turn depends on several factors:

- If the contact between the trace originator and the tracee occurred environmentally (determined with probability *p*_*env*_), tracing cannot take place.
- If transmission was not environmental, the contact can be traced digitally if:
  ○ Both the trace originator and the tracee possess chirp-enabled smartphones (determined independently for each individual case with probability *p*_*smartphone*_);
  ○ The trace originator shares their data with the tracing app (determined independently for each individual case with probability *p*_*share*_*digital*_);
  ○ The time of between contact (equal to the exposure time of the child case, i.e. of the trace in forward tracing and the trace initiator in reverse tracing) and trace initiation is less than the **data-retention window** of the digital tracing system;
  ○ The contact between the two cases was recorded by the tracing app of the trace originator (determined independently for each individual case with probability *p*_*trace*_*digital*_).
- If any of the above conditions are not met, but transmission was not environmental, the contact might still be traced manually if:
  ○ The trace originator shares their contact history with a manual contact tracer (determined independently for each individual case with probability *p*_*share*_*manual*_);
  ○ The time between contact (as above) and the identification time or symptom onset of the trace initiator (whichever came first) is less than the **contact-tracing window** of the manual tracing system;
  ○ The tracee is successfully traced by the contact tracer (determined independently for each individual case with probability *p*_*trace*_*manual*_).
- If neither digital nor manual tracing succeeds, then the trace fails and the tracee is not traced.

Cases that are successfully traced are identified at a time equal to the **trace initiation time** of the trace originator plus a delay time drawn from the appropriate **trace delay distribution** (which will differ between digital and manual tracing). Cases identified through tracing can then be isolated, tested, and traced as described above. If a case is isolated through tracing earlier than they would have been otherwise, child cases whose exposure time would be later than their parent’s new isolation time are eliminated, as are their descendents.

### Run initiation and termination

A simulation of an outbreak under the branching-process model is initialised with a given number of index cases (by default 20, in order to reduce the probability of stochastic elimination) and proceeds generation by generation until either no further child cases are generated (extinction) or the run exceeds one of:

1. A **cumulative case limit** of 10,000 cases, reached if the total number of cases ever exceeded that number, or
2. A **time limit** of 52 weeks, reached if the latest exposure time across all cases ever exceeded that number.

In practice, virtually all runs either went extinct or reached the cumulative case limit; across all scenarios tested for all datasets used in Figures 2-4, the overall percentage of runs that terminated as a result of exceeding the time limit was less than 0.02%, and the highest percentage observed for any single scenario was 1.3%. The cumulative case limit, meanwhile, was selected to minimise the chance of a run that would otherwise go extinct being terminated prematurely while preserving computational tractability; in test runs with a cumulative case limit of 100,000 cases, fewer than 2% of extinct runs in any scenario had a cumulative case count of over 10,000.

A terminated run was deemed “controlled” if it reached extinction, and uncontrolled otherwise. The control rate for a scenario was computed as the proportion of runs for that scenario that were controlled. 95% credible intervals on the control rate were computed by beta-binomial conjugacy under a *Beta*(1, 1) uniform prior, as the 2.5th and 97.5th percentiles of the beta distribution *Beta*(1 + *k*, 1 + *n* − *k*), where *n* is the total number of runs for that scenario and *k* is the number of controlled runs. Effective reproduction numbers were computed as the average number of child cases produced across all cases in a run, averaged across all runs in the scenario. For main figures, 1000 runs were performed per scenario; for figures, either 500 or 1000 runs were performed, as specified in the figure captions.

## 2. Supplementary tables

**Table S1:** Parameters of the branching-process model.

| Parameter | Value |  |  | Sources and Notes |
| --- | --- | --- | --- | --- |
|  | Median | Optimistic | Pessimistic |  |
| % asymptomatic carriers | 45% | 40% | 55% | 8–10,31 |
| Relative infectiousness of asymptomatic carriers | 50% | 45% | 60% | Informed by viral loads and tracing results described in <sup>5,9,23,25,26,32</sup> |
| % environmental transmission | 10% | 5% | 15% | 4,33 |
| Proportion of pre-symptomatic transmission | 48% | 38% | 53% | Informed by viral load measurements, tracing results, and negative serial intervals described in <sup>5,23,26,34–39</sup> |
| % of symptomatic cases identified without tracing | 90% |  |  | Assumed |
| % of cases with chirping smartphones (high-uptake case) | 53% (low-uptake) / 80% (high-uptake) |  |  | Survey data on phone ownership and attitudes to exposure notification<sup>19,20,40</sup> |
| % of cases with smartphones who upload data when diagnosed | 90% |  |  | Assumed |
| % of cases who share data with manual contact tracers | 98% |  |  | Non-cooperation reportedly rare<sup>41</sup> |
| Test sensitivity | 80% |  |  | 42,43 |
| $R_0$ (default) | 2.5 | | | Most estimates cluster between 2.0 and 3.0: <sup>4,9,22–24,44</sup> |
| Overdispersion | 0.11 |  |  | 45 |
| Number of initial cases | 20 |  |  | Assumed |
| Data retention window for digital tracing (days) | 14 days |  |  | 46 |
| Incubation period | 5.5 ± 2.1 days (lognormal distribution) |  |  | 47 |
| Delay from onset to isolation | 3.8 ± 2.4 days (Weibull distribution) |  |  | 17 |
| Delay for testing | 1 ± 0.3 days (gamma distribution) |  |  | Assumed |
| Delay for manual tracing | 1.5 ± 4.8 days (lognormal distribution); median 0.5 days |  |  | Previous reports suggest most contacts can be traced within one day, but some take much longer<sup>48</sup> |
| Delay for digital tracing | 0 days |  |  | Assumed |

## 3. Supplementary figures

**Figure S1:**
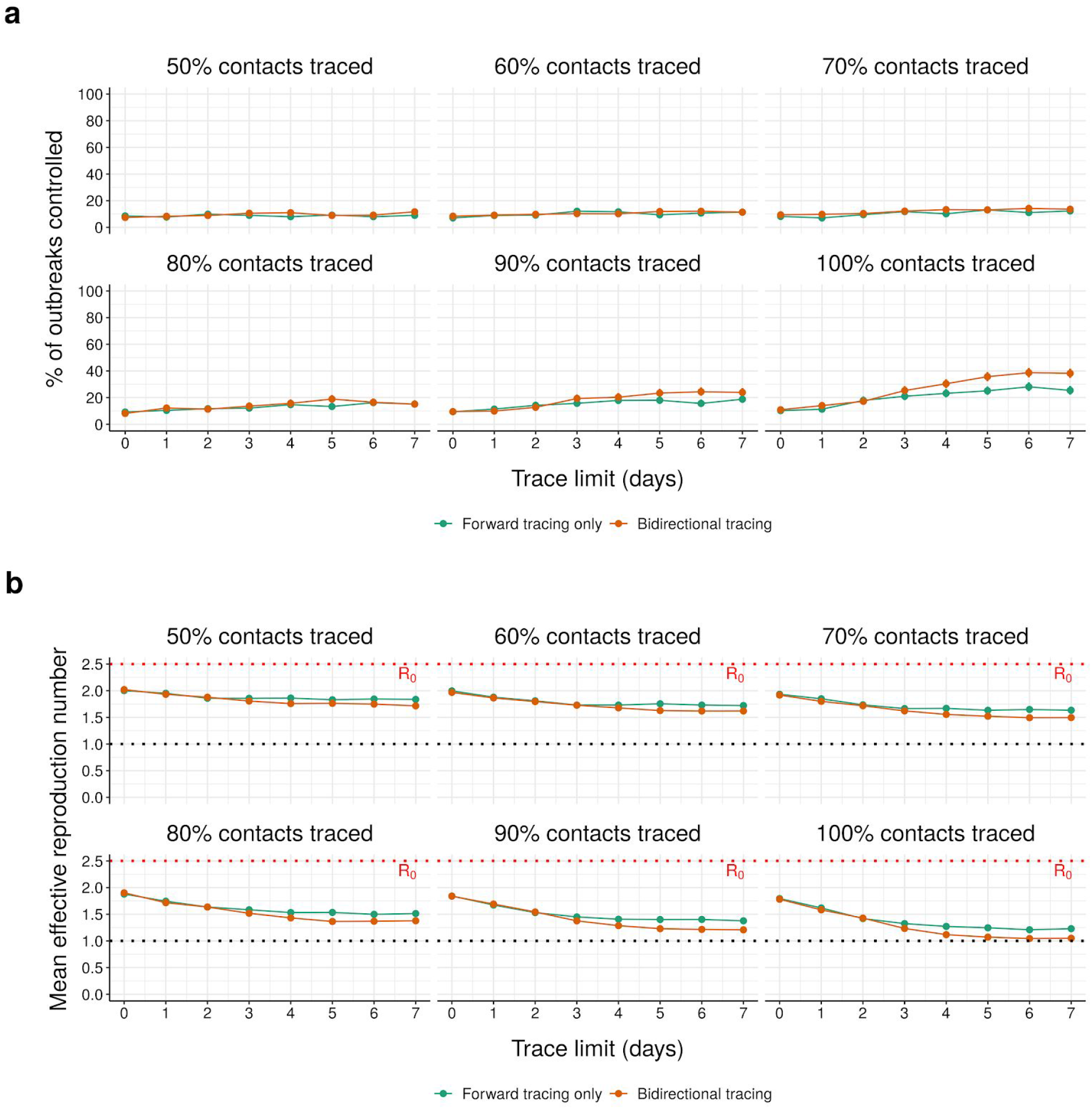
Effect of contact-tracing window size on performance of manual tracing. (a) Control rate and (b) average effective reproduction number of manual tracing under median parameters (Table S1) when contacts are traced for varying periods prior to symptom onset (for symptomatic cases) or case identification (for presymptomatic and asymptomatic cases). Panel headers indicate the percentage of non-environmental contacts traced. Error bars in (a) represent 95% credible intervals across 1000 runs under a uniform beta prior; points in (b) represent average values over the same.

**Figure S2:**
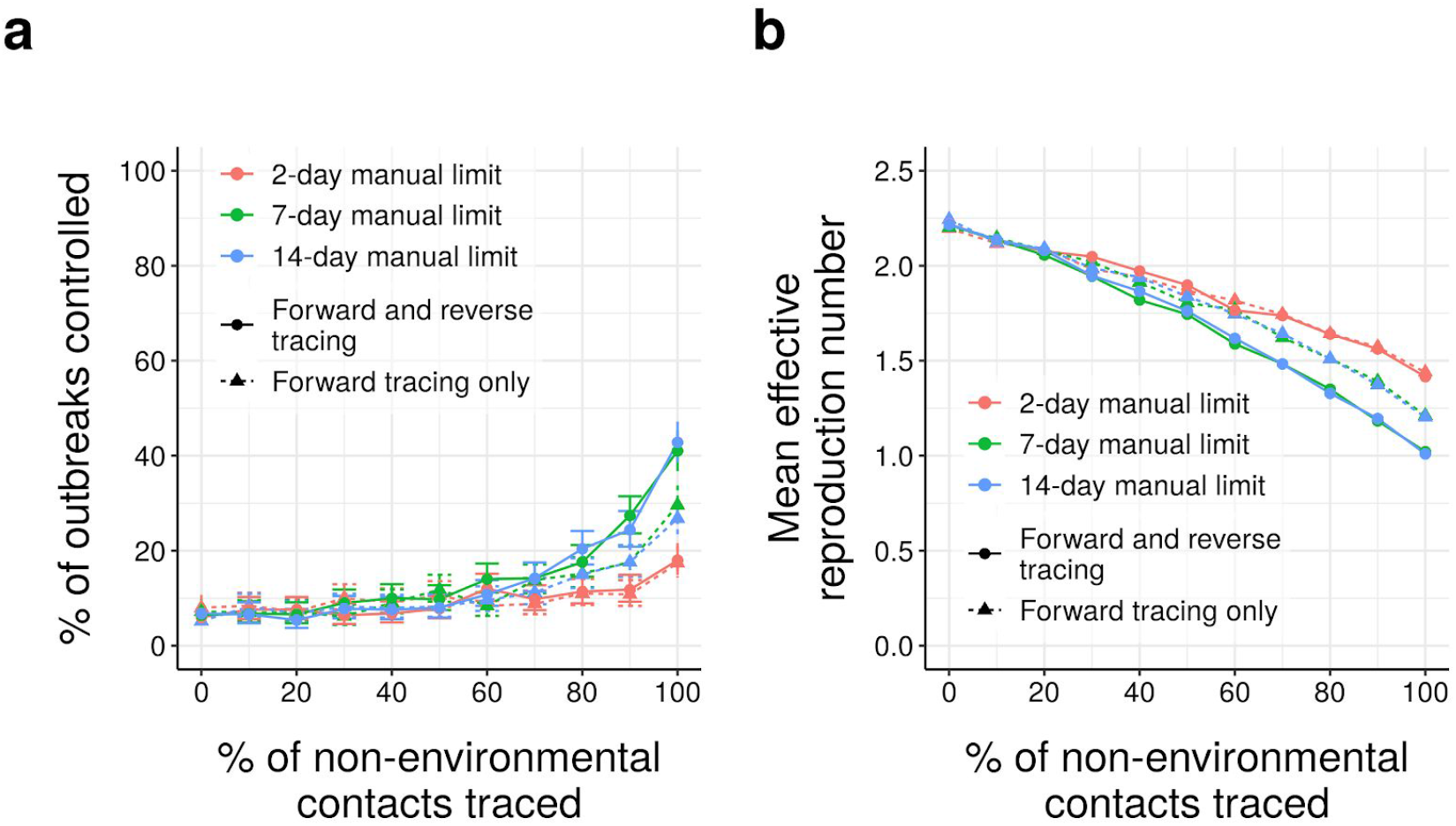
Effect of increased contact-tracing window size on performance of manual tracing. (a) Control rate and (b) average effective reproduction number of manual tracing under median disease parameters (Table S1) when contacts are traced up to 2, 7, or 14 days prior to symptom onset (for symptomatic cases) or case identification (for presymptomatic and asymptomatic cases). Error bars in (a) represent 95% credible intervals across 500 runs under a uniform beta prior; points in (b) represent average values over the same.

**Figure S3:**
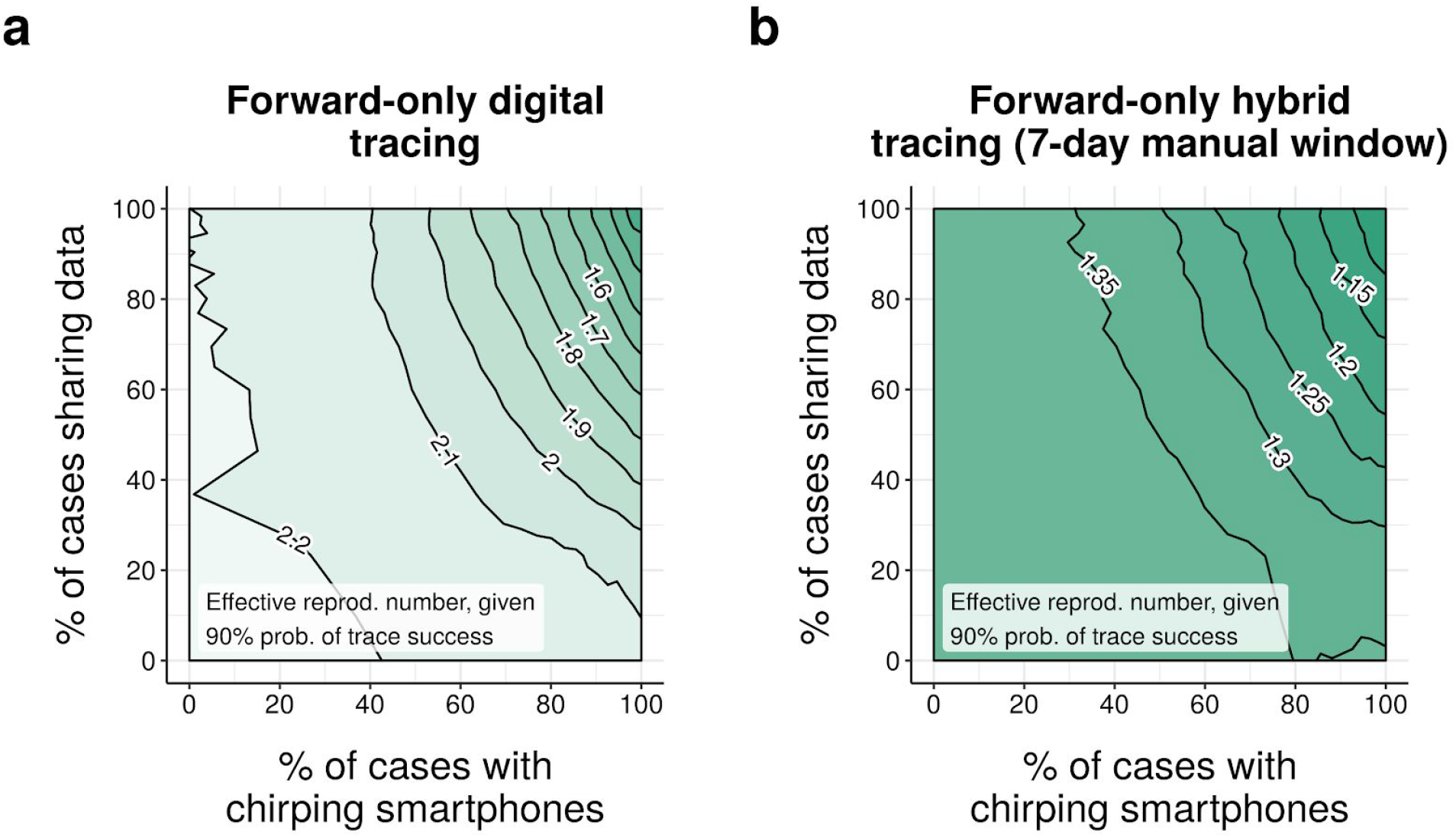
Effect of network fragmentation on performance of forward-only tracing. Neighbour-averaged contour plot of effective reproduction number achieved under (a) digital-only and (b) hybrid forward-only tracing, over 1000 runs per scenario, for different levels of smartphone coverage and data-sharing, assuming median disease parameters (Table S1) and a 90% probability of trace success.

**Figure S4:**
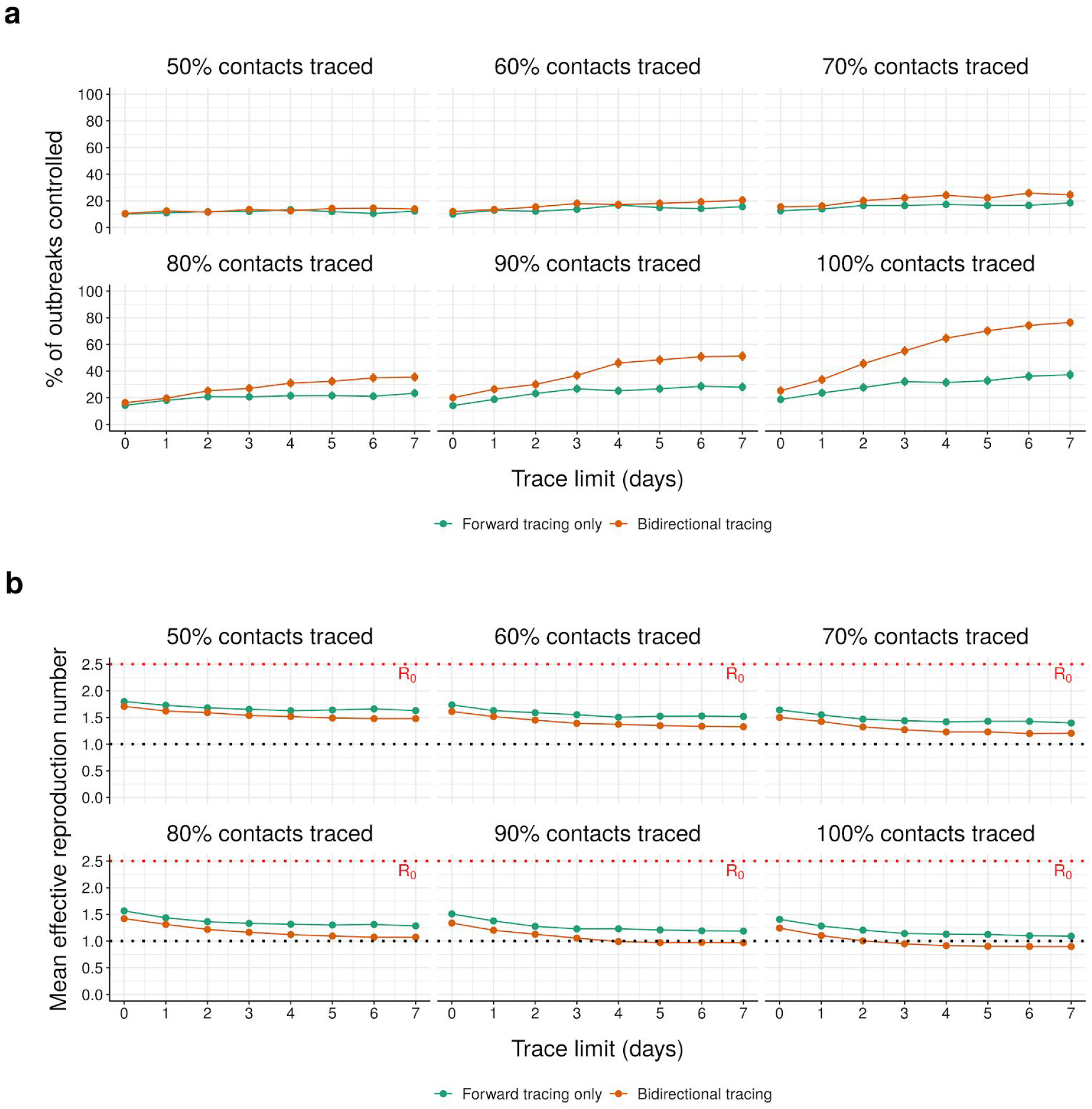
Effect of contact-tracing window size on performance of hybrid tracing. (a) Control rate and (b) average effective reproduction number of hybrid tracing under median parameters (Table S1) when contacts are manually traced for varying periods prior to symptom onset (for symptomatic cases) or case identification (for presymptomatic and asymptomatic cases). Data for digital contact tracing is assumed to be retained for 14 days after contact. Panel headers indicate the percentage of non-environmental contacts traced. Error bars in (a) represent 95% credible intervals across 1000 runs under a uniform beta prior; points in (b) represent average values over the same.

**Figure S5:**
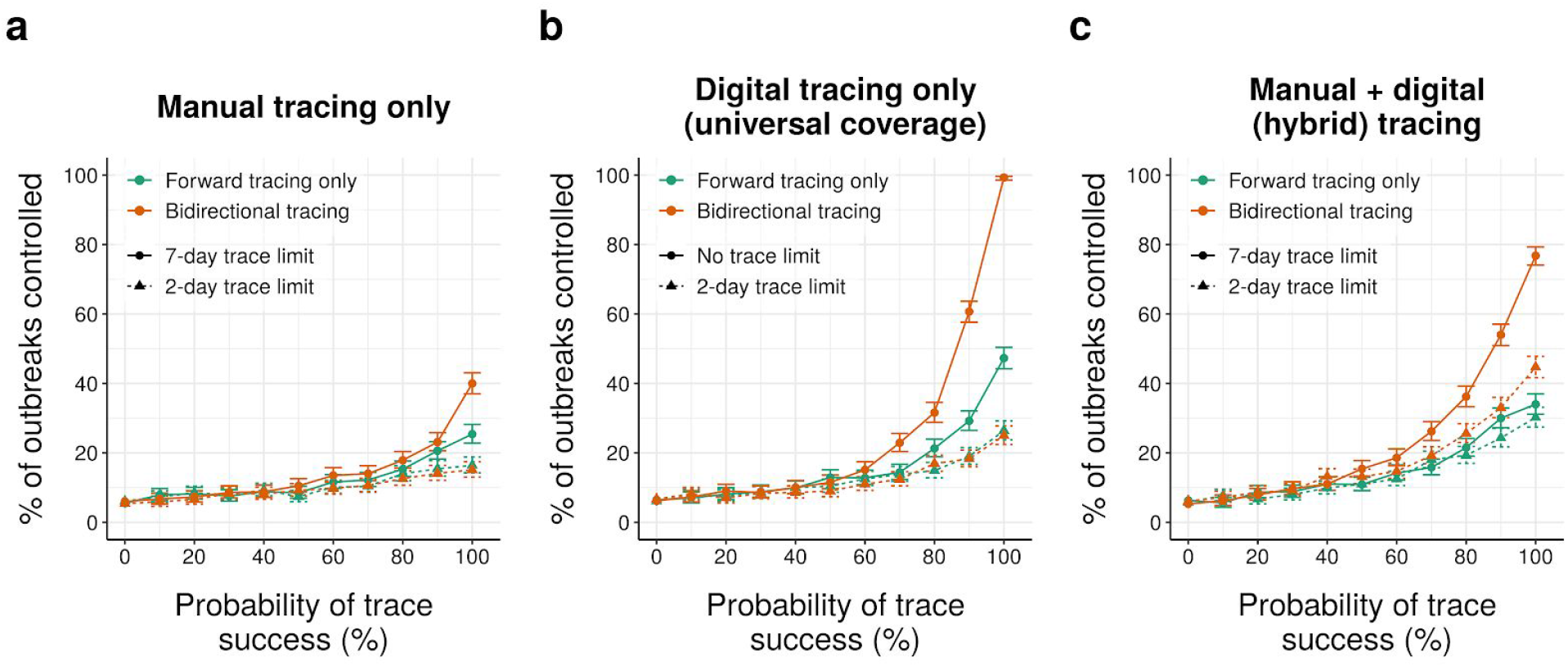
Outbreak control under manual, digital and hybrid tracing tracing. As Fig. 2a,b,d, but showing % outbreaks controlled rather than effective reproduction number. Points represent average values over 1000 runs.

**Figure S6:**
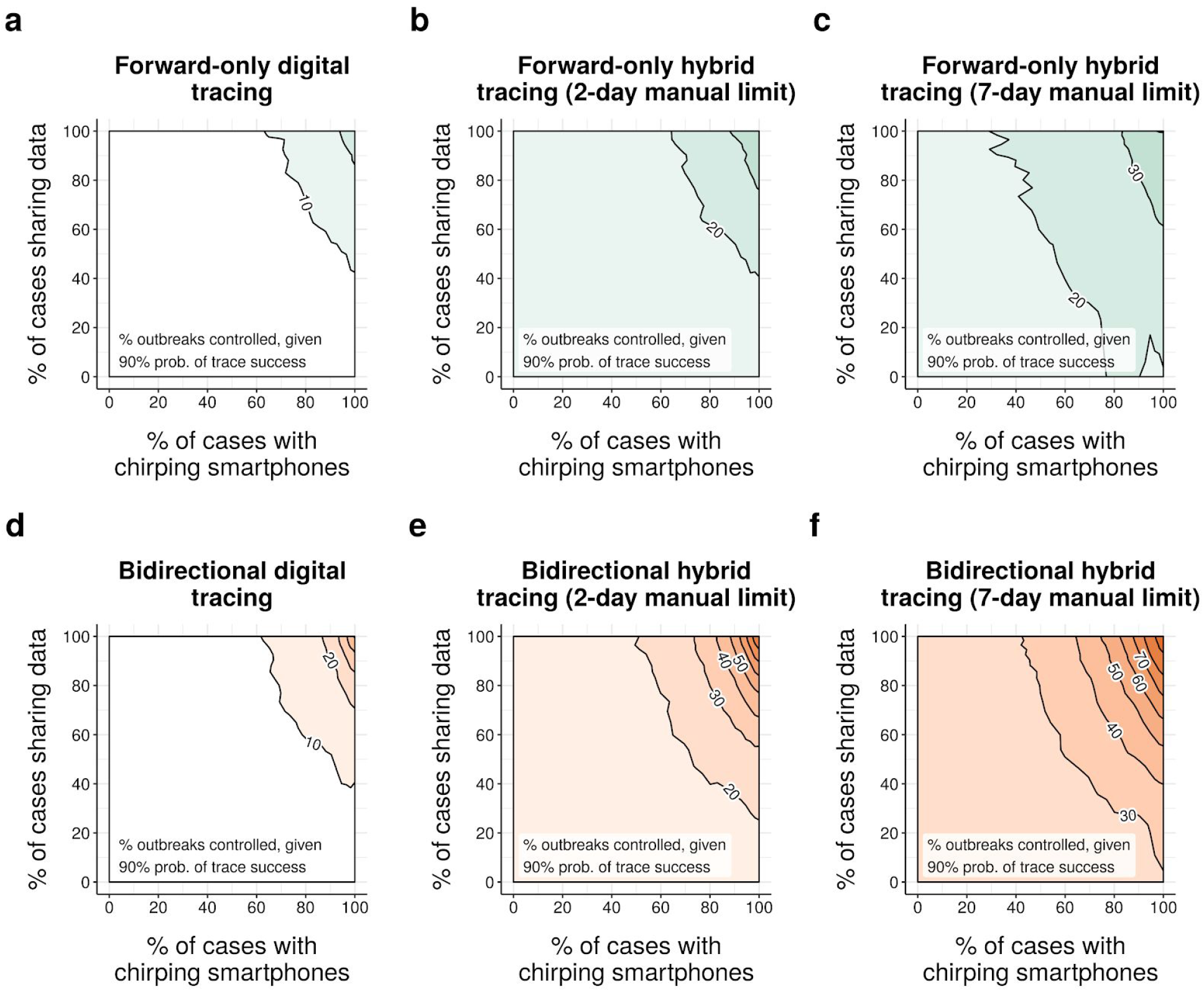
Effect of network fragmentation on effective reproduction number (% outbreaks controlled). Neighbour-averaged contour plots of % outbreaks controlled (over 1000 runs per scenario) under (a-c) forward-only and (d-f) bidirectional tracing, assuming median disease parameters (Table S1) and a 90% probability of trace success.

**Figure S7:**
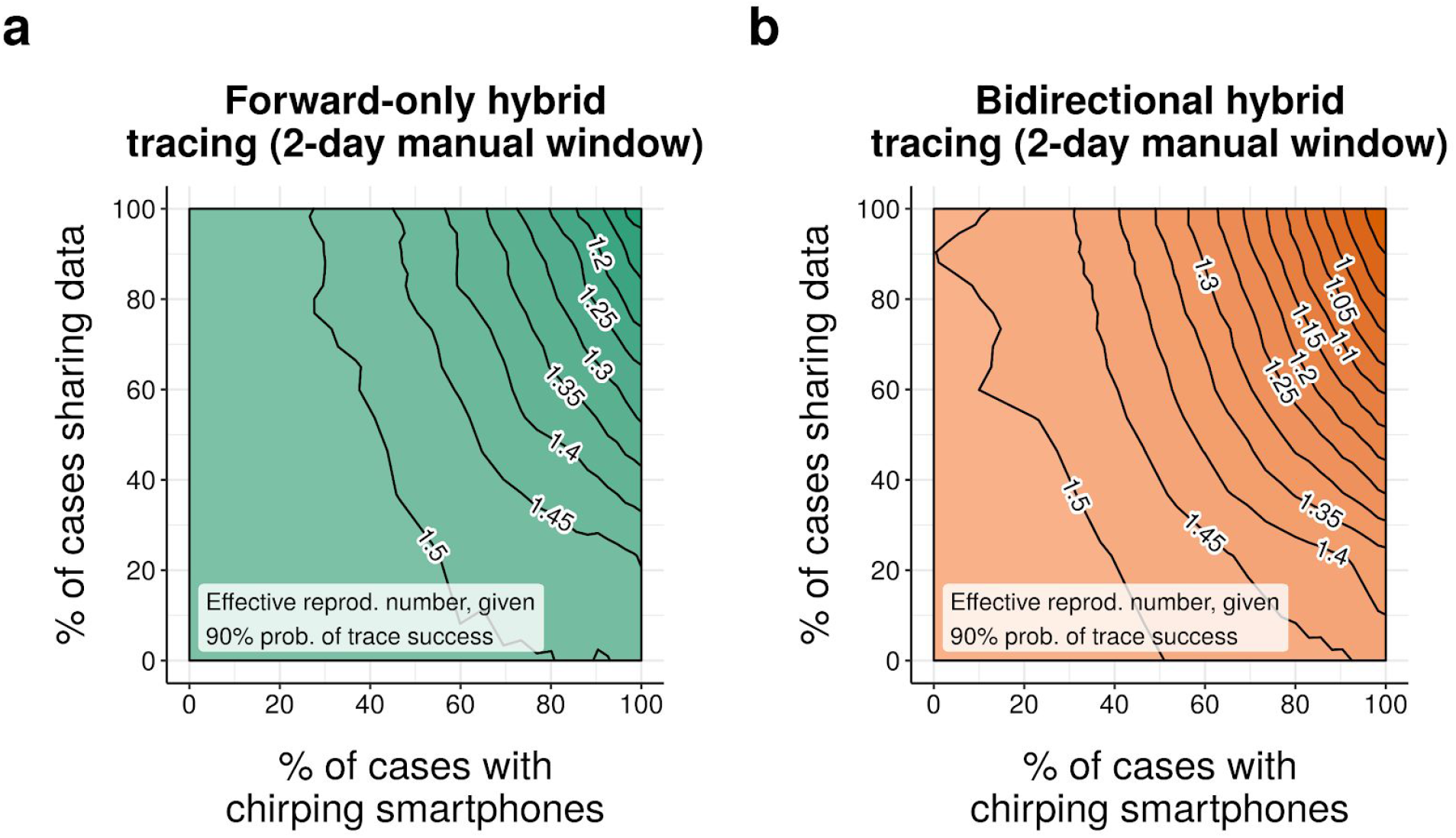
Effect of network fragmentation on control rates under a 2-day manual tracing window. Neighbour-averaged contour plots of effective reproduction number achieved (over 1000 runs per scenario) under (a) bidirectional and (b) forward-only hybrid (manual+digital) tracing with a 2-day manual tracing window, assuming median disease parameters (Table S1) and a 90% probability of trace success.

**Figure S8:**
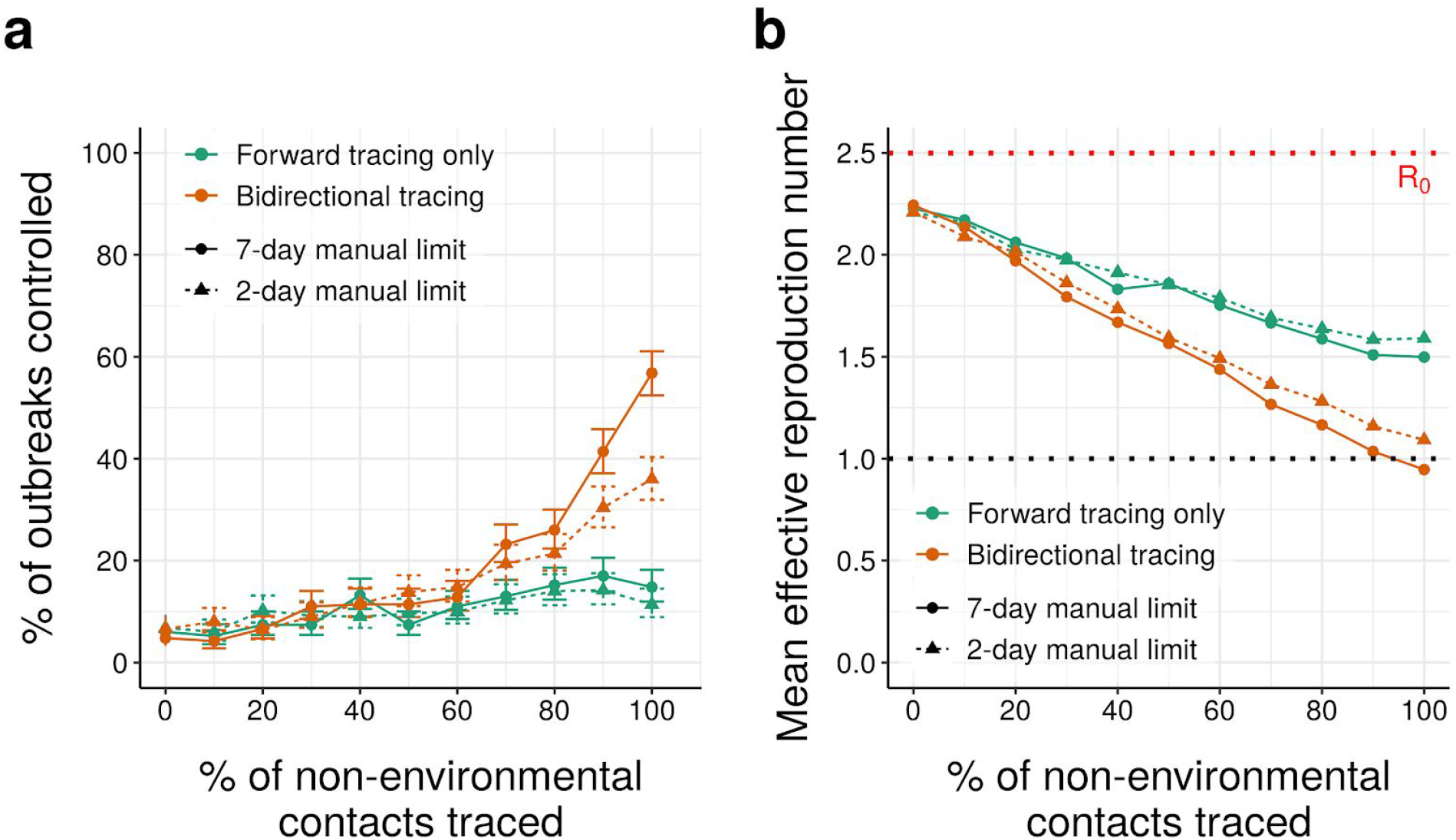
Effect of requiring pre-trace testing on hybrid tracing performance. As Fig. 3e-f, but requiring a positive test result before initiating contact tracing from a symptomatic case. Error bars in (a) represent 95% credible intervals across 500 runs under a uniform beta prior; points in (b) represent average values over the same.

**Figure S9:**
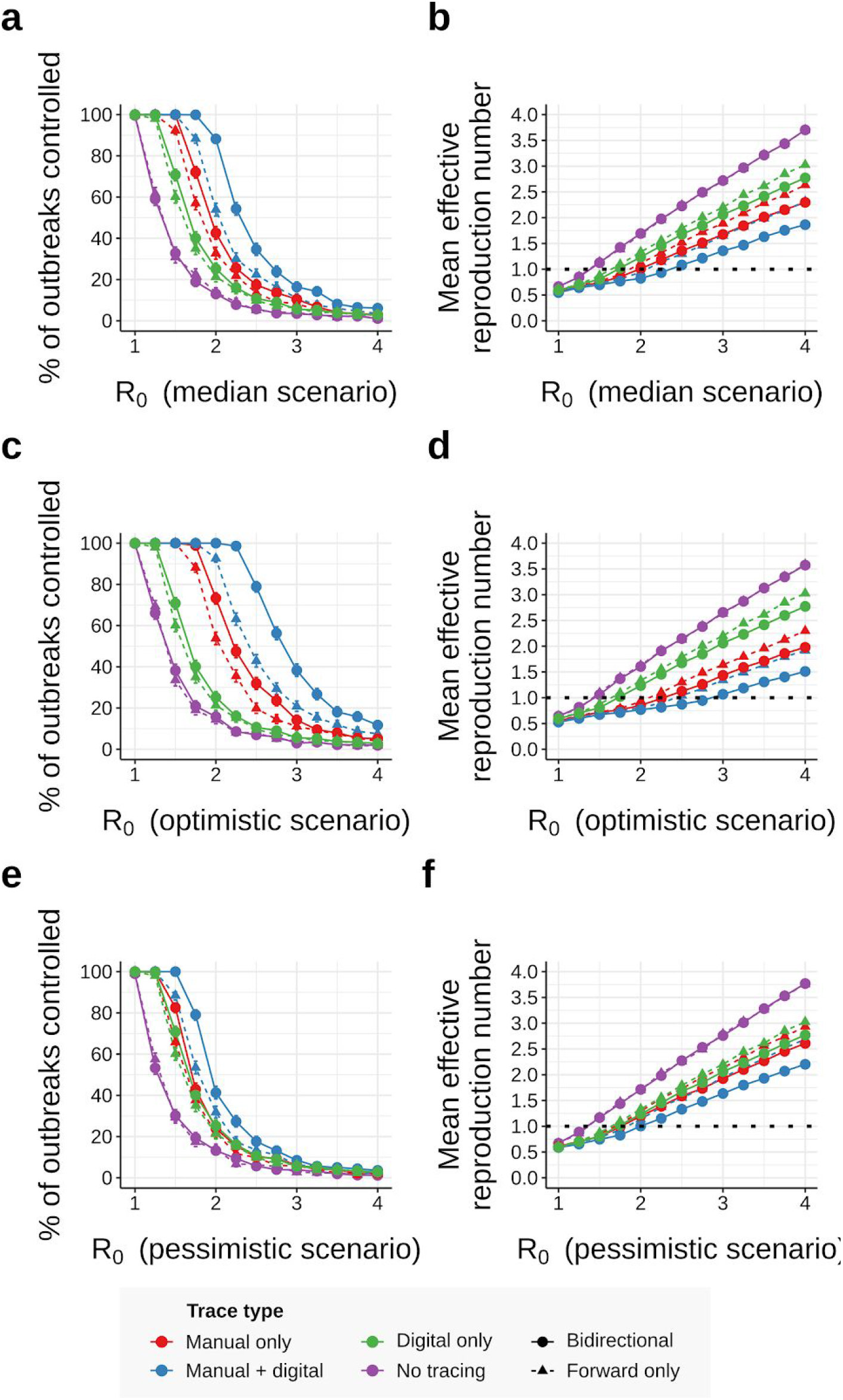
Effect of R_0_ and disease parameters on tracing performance (80% trace rate). As Figure 3, but assuming 80% of non-environmental contacts are traced. Error bars in (a,c,e) represent 95% credible intervals across 1000 runs under a uniform beta prior; points in (b) represent average values over the same.

**Figure S10:**
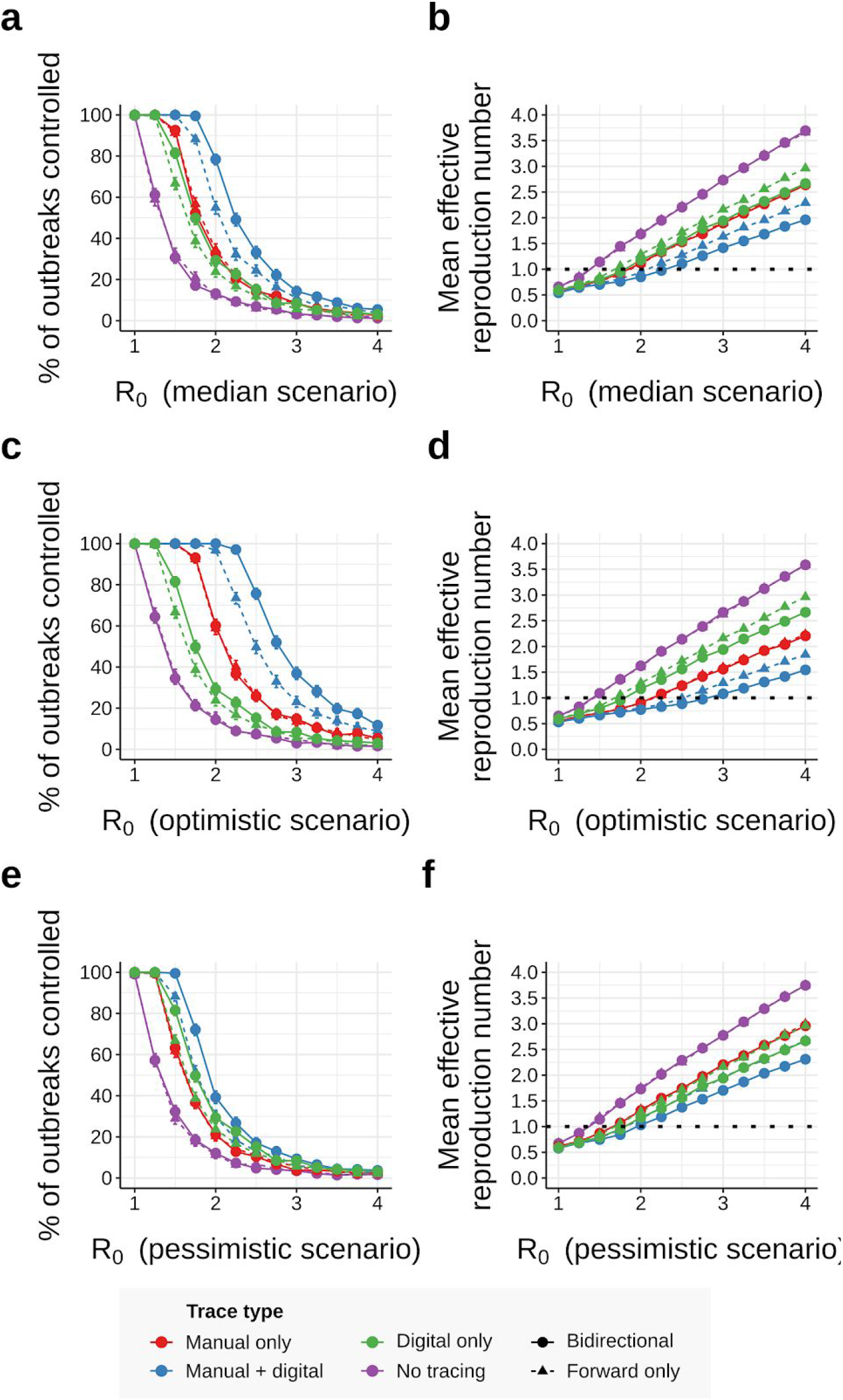
Effect of R_0_ and disease parameters on tracing performance (2-day manual trace window). As Figure 3, but assuming a 2-day manual trace window. Error bars in (a,c,e) represent 95% credible intervals across 1000 runs under a uniform beta prior; points in (b,d,f) represent average values over the same.

**Figure S11:**
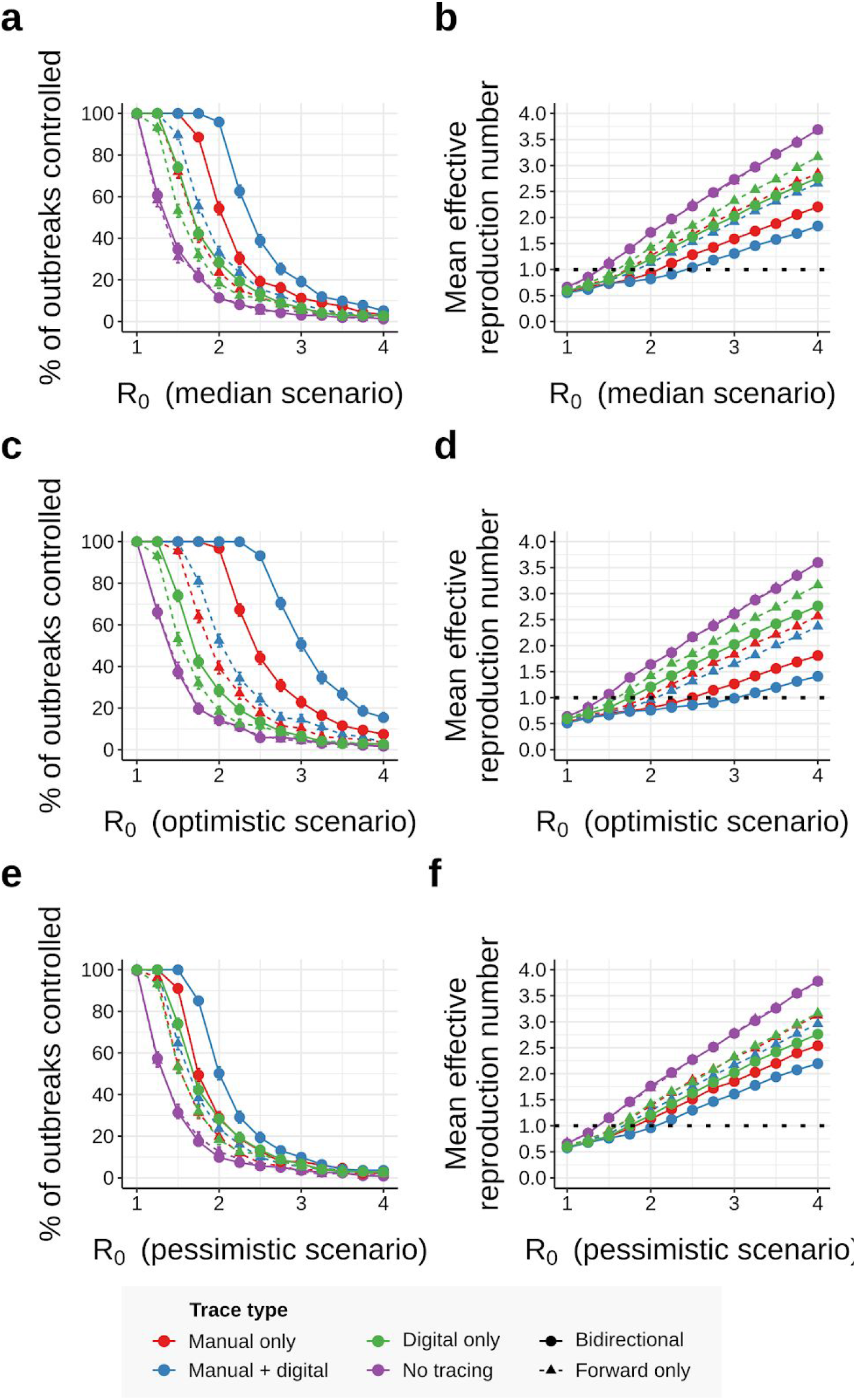
Effect of R_0_ and disease parameters on tracing performance (pre-emptive testing). As Figure 3, but requiring a positive test result before tracing symptomatic cases. Error bars in (a,c,e) represent 95% credible intervals across 1000 runs under a uniform beta prior; points in (b,d,f) represent average values over the same.

**Figure S12:**
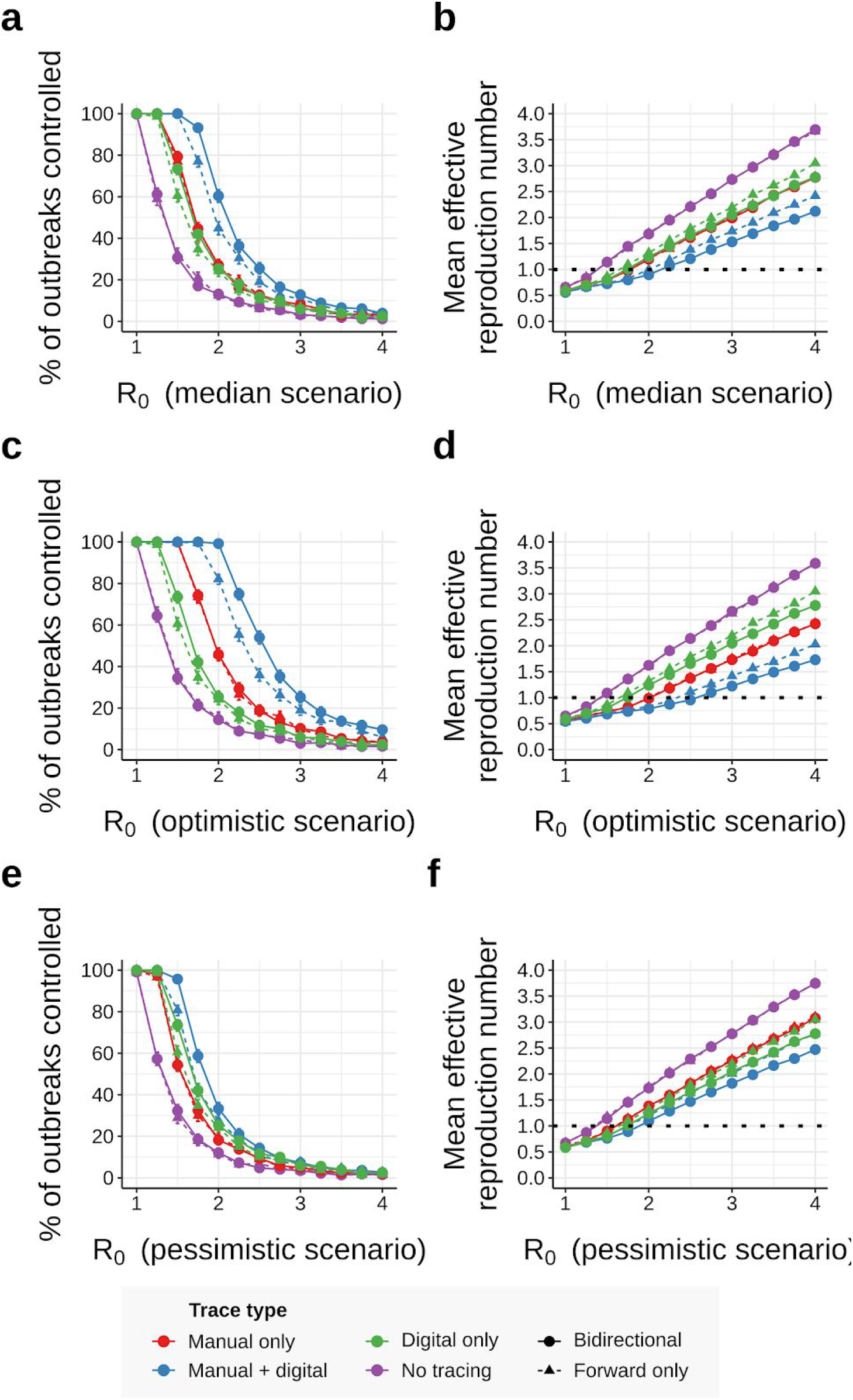
Effect of R_0_ and disease parameters on tracing performance (80% trace rate, 2-day manual trace window). As Figure 3, but assuming 80% of non-environmental contacts traced and a 2-day manual trace window. Error bars in (a,c,e) represent 95% credible intervals across 1000 runs under a uniform beta prior; points in (b,d,f) represent average values over the same.

**Figure S13:**
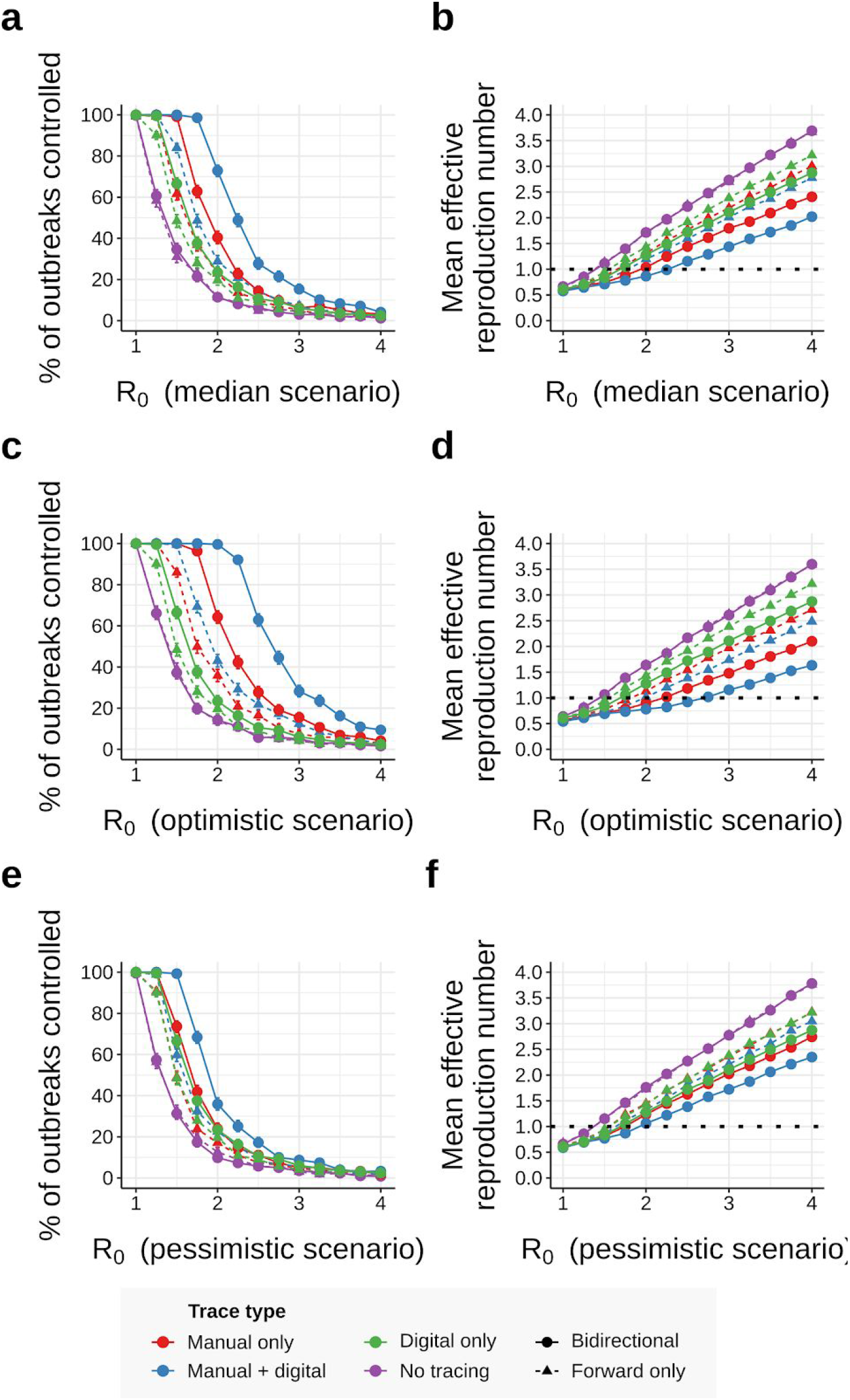
Effect of R_0_ and disease parameters on tracing performance (pre-emptive testing, 80% trace rate). As Figure 3, but requiring a positive test result before tracing symptomatic cases and assuming 80% of non-environmental contacts traced and a 2-day manual trace window. Error bars in (a,c,e) represent 95% credible intervals across 1000 runs under a uniform beta prior; points in (b,d,f) represent average values over the same.

**Figure S14:**
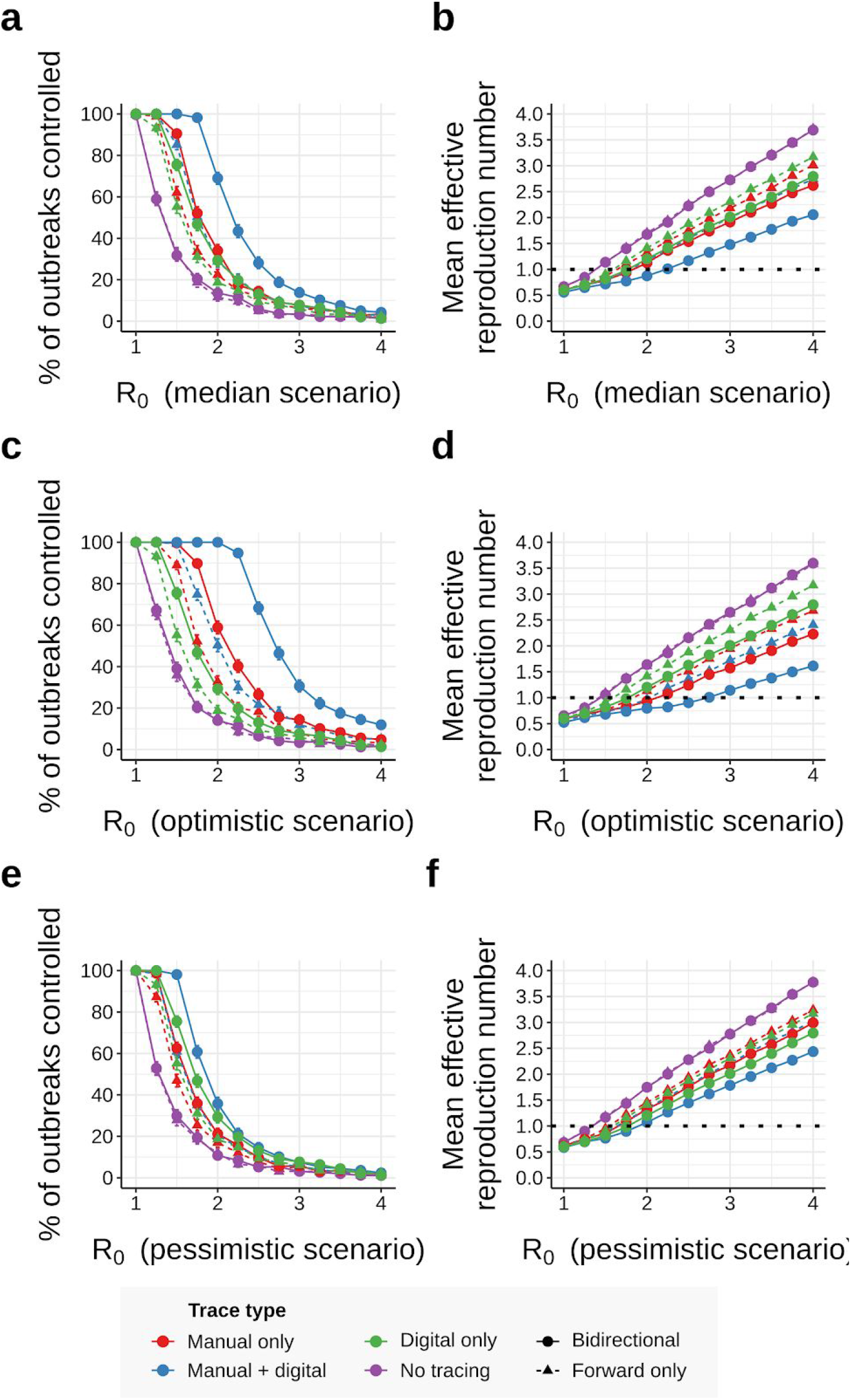
Effect of R_0_ and disease parameters on tracing performance (pre-emptive testing, 2-day manual trace window). As Figure 3, but requiring a positive test result before tracing symptomatic cases and assuming a 2-day manual trace window. Error bars in (a,c,e) represent 95% credible intervals across 1000 runs under a uniform beta prior; points in (b,d,f) represent average values over the same.

**Figure S15:**
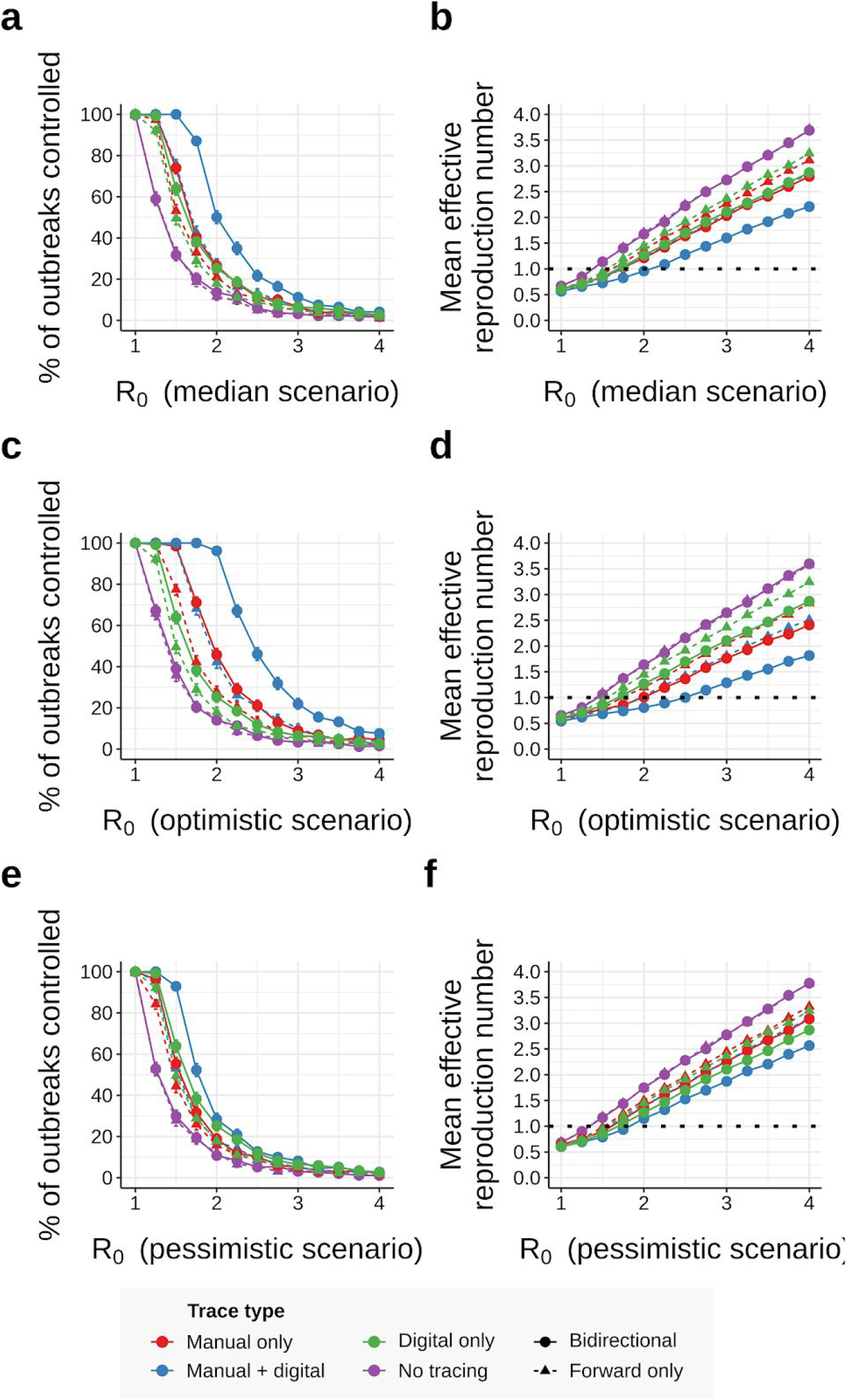
Effect of R_0_ and disease parameters on tracing performance (pre-emptive testing, 80% trace rate, 2-day manual trace window). As Figure 3, but requiring a positive test result before tracing symptomatic cases, and assuming 80% of non-environmental contacts traced and a 2-day manual trace window. Error bars in (a,c,e) represent 95% credible intervals across 1000 runs under a uniform beta prior; points in (b,d,f) represent average values over the same.

**Figure S16:**
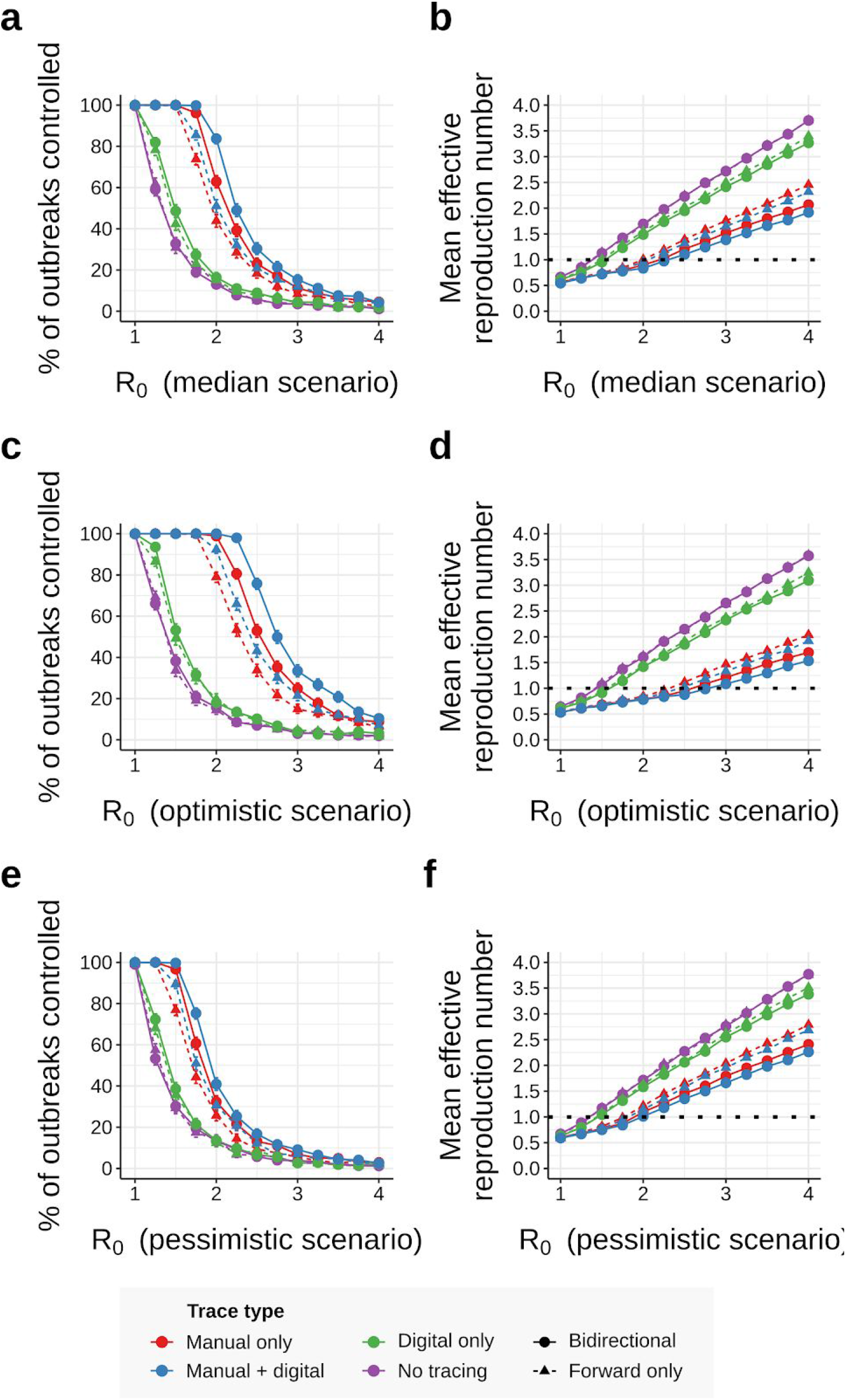
Effect of R_0_ and disease parameters on tracing performance (low-uptake case). As Figure 3, but assuming only 53% of cases have chirp-enabled smartphones. Error bars in (a,c,e) represent 95% credible intervals across 1000 runs under a uniform beta prior; points in (b,d,f) represent average values over the same.

**Figure S17:**
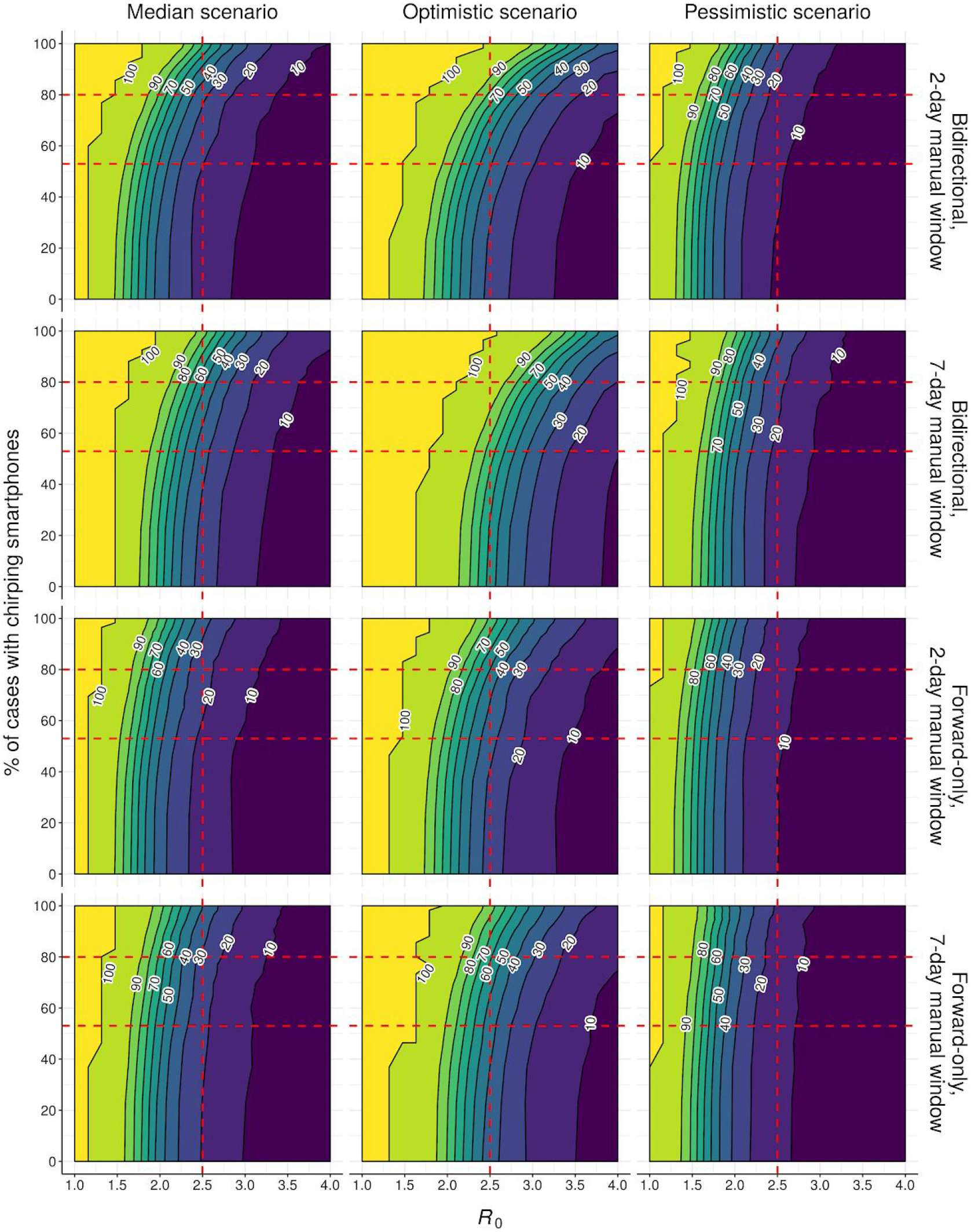
Effect of R0, network fragmentation, and disease parameters on hybrid tracing performance. Neighbour-averaged contour plots showing % of outbreaks controlled (over 500 runs) under hybrid tracing for different disease scenarios (Table S1) and tracing strategies, assuming 90% of non-environmental contacts traced and immediate tracing of symptomatic cases. Horizontal red dashed lines indicate low- and high-uptake scenarios from the main text (Table S1); vertical red dashed line indicates the *R*_0_ value used in Fig. 2 & 4.

**Figure S18:**
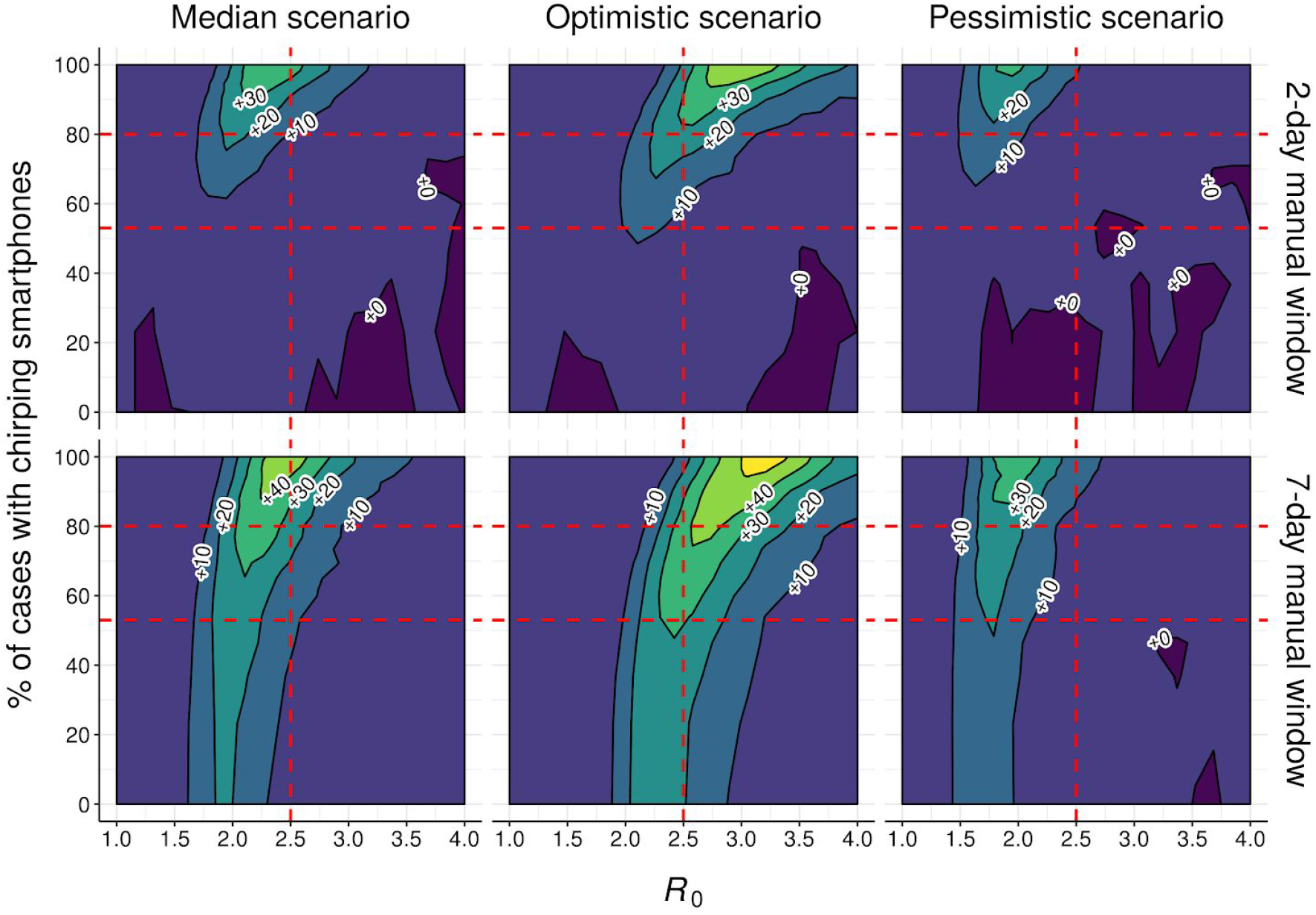
Effect of R0, network fragmentation, and disease parameters on outperformance of bidirectional tracing. Contour plot showing difference in % of outbreaks controlled (over 500 runs) between bidirectional and forward-only hybrid tracing for different disease scenarios (Table S1) and tracing strategies, assuming 90% of non-environmental contacts traced and immediate tracing of symptomatic cases. Horizontal red dashed lines indicate low- and high-uptake scenarios from the main text (Table S1); vertical red dashed line indicates the *R*_0_ value used in Fig. 2 & 4.

**Figure S19:**
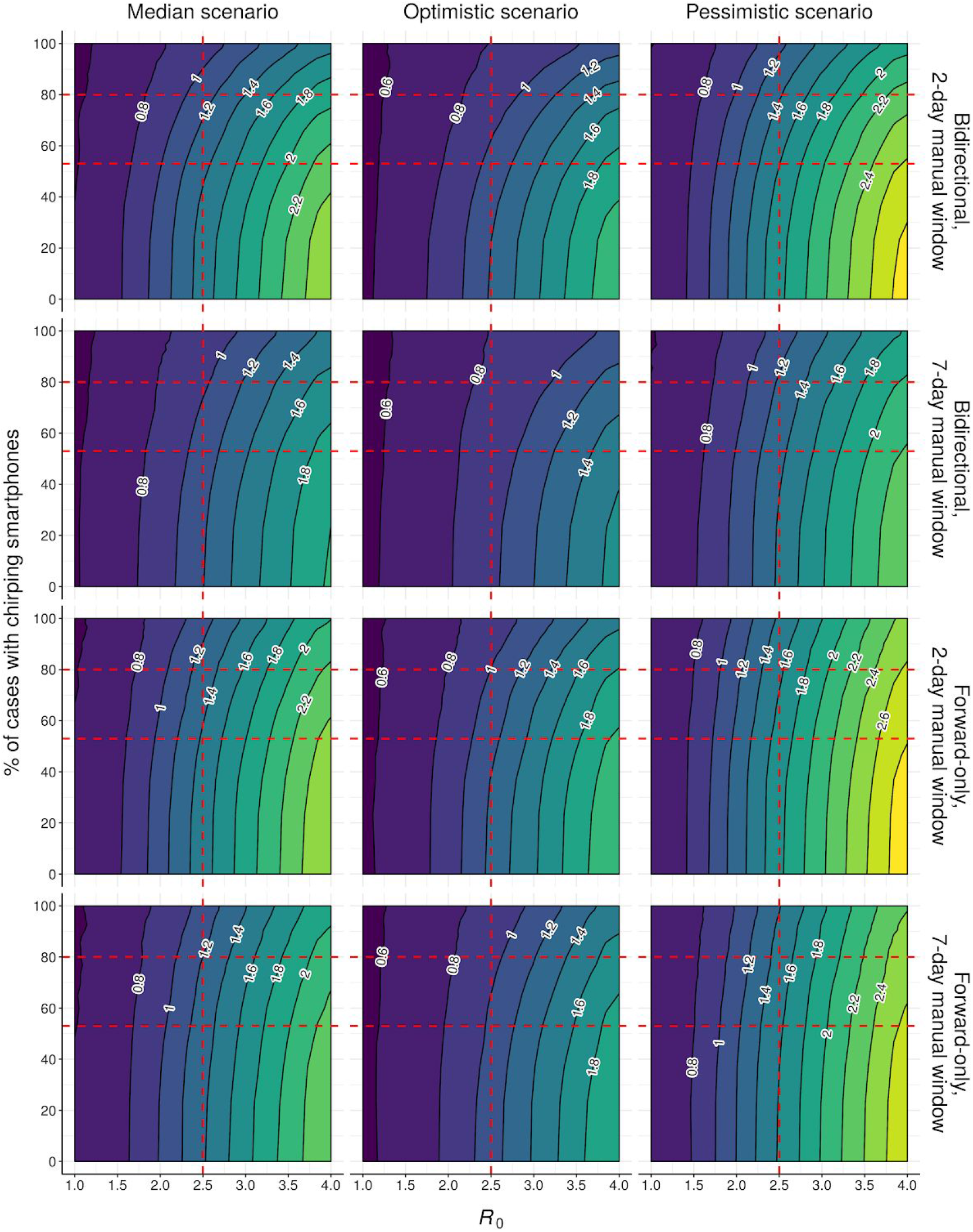
Effect of R0, network fragmentation, and disease parameters on hybrid tracing performance (R_eff_). Contour plot showing mean effective reproduction number (over 500 runs) under hybrid tracing for different disease scenarios (Table S1) and tracing strategies, assuming 90% of non-environmental contacts traced and immediate tracing of symptomatic cases. Horizontal red dashed lines indicate low- and high-uptake scenarios from the main text (Table S1); vertical red dashed line indicates the *R*_0_ value used in Fig. 2 & 4.

**Figure S20:**
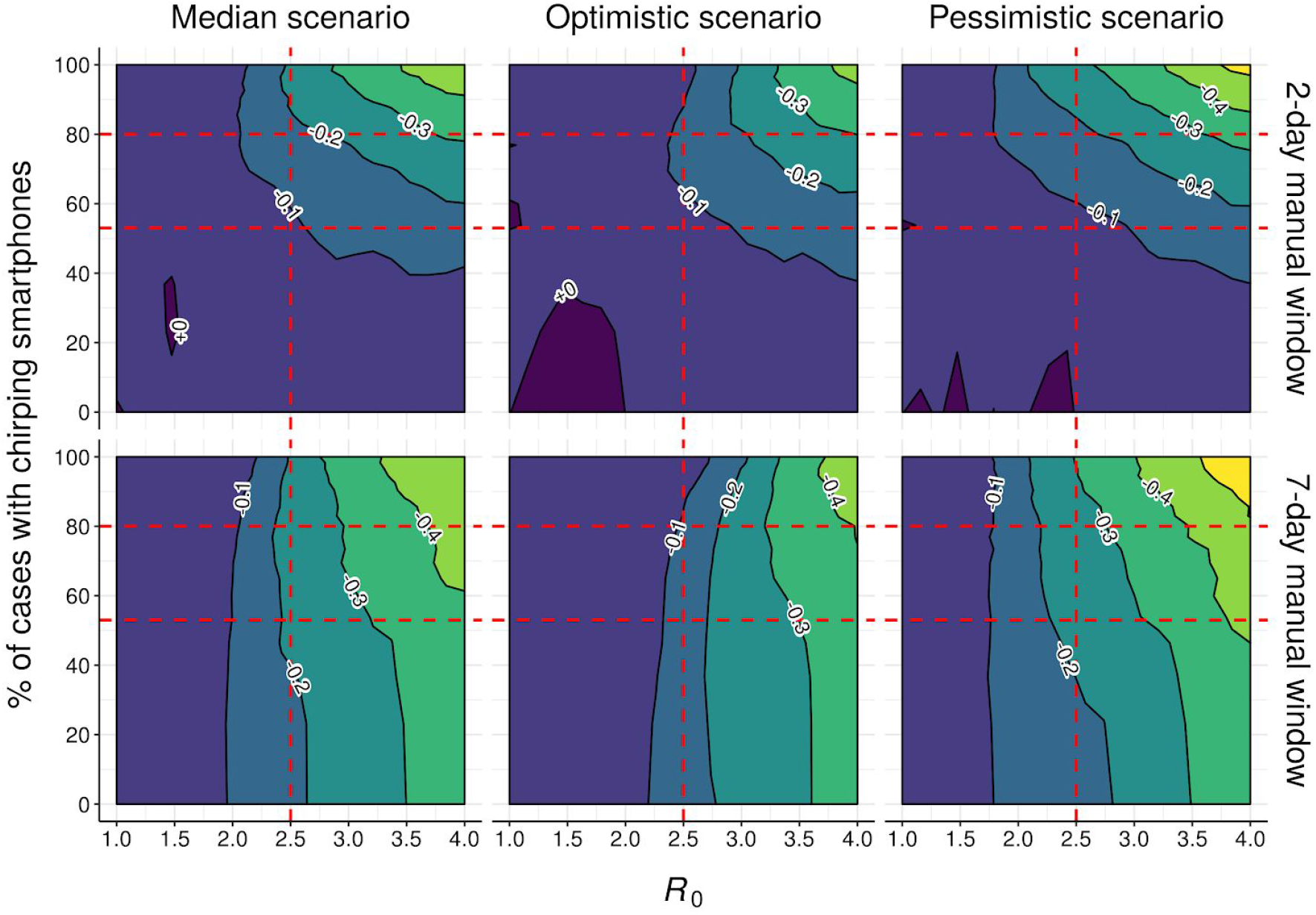
Effect of R0, network fragmentation, and disease parameters on outperformance of bidirectional tracing (R_eff_). Contour plot showing difference in mean effective reproduction number (over 500 runs) between bidirectional and forward-only hybrid tracing for different disease scenarios (Table S1) and tracing strategies, assuming 90% of non-environmental contacts traced and immediate tracing of symptomatic cases. Horizontal red dashed lines indicate low- and high-uptake scenarios from the main text (Table S1); vertical red dashed line indicates the *R*_0_ value used in Fig. 2 & 4.

**Figure S21:**
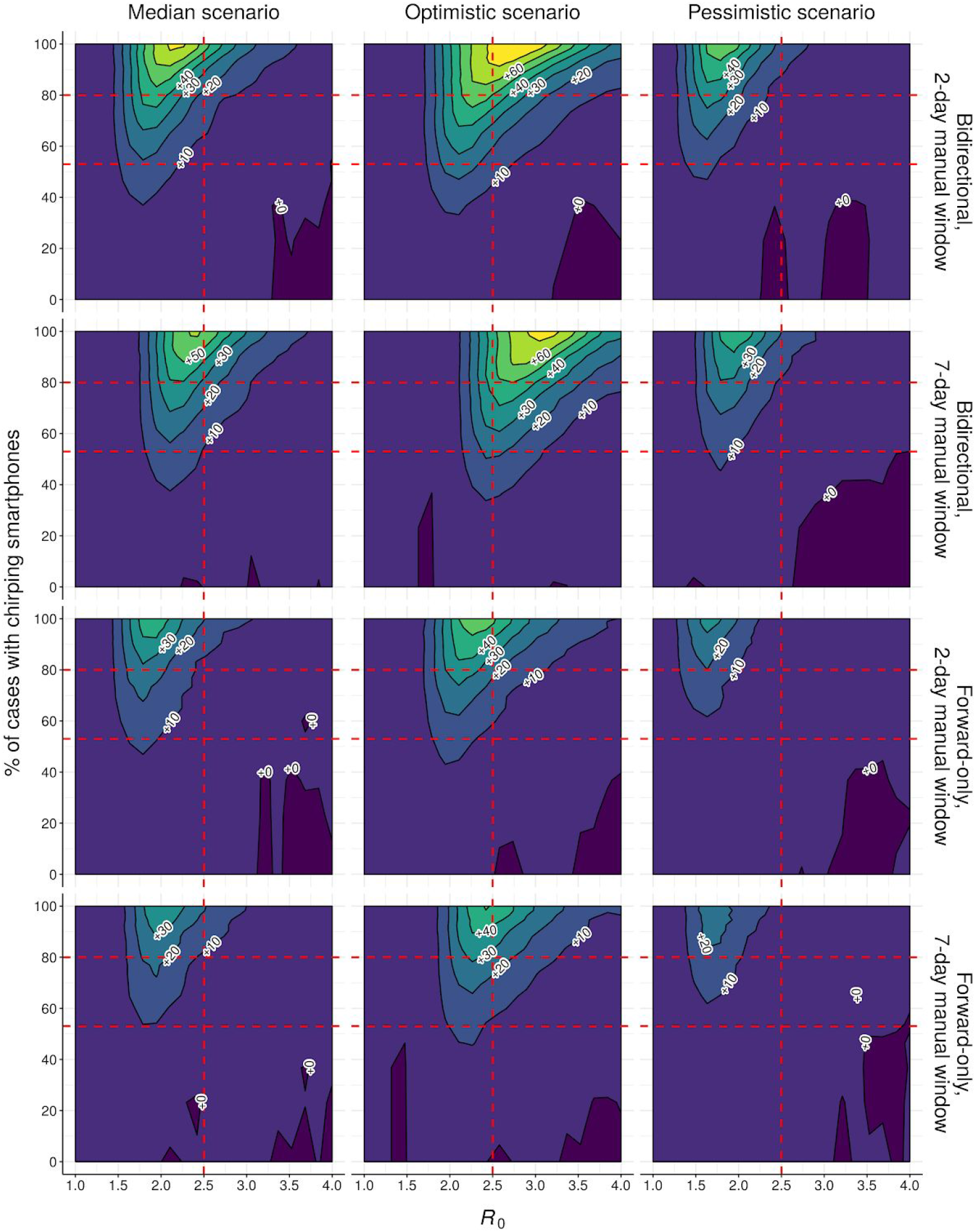
Effect of R0, network fragmentation, and disease parameters on outperformance of hybrid vs manual tracing. Contour plot showing difference in % outbreaks controlled (over 500 runs) between hybrid and manual-only hybrid tracing for different disease scenarios (Table S1) and tracing strategies, assuming 90% of non-environmental contacts traced and immediate tracing of symptomatic cases. Horizontal red dashed lines indicate low- and high-uptake scenarios from the main text (Table S1); vertical red dashed line indicates the *R*_0_ value used in Fig. 2 & 4. Note that the data-retention limit for the digital system is 14 days in all cases.

**Figure S22:**
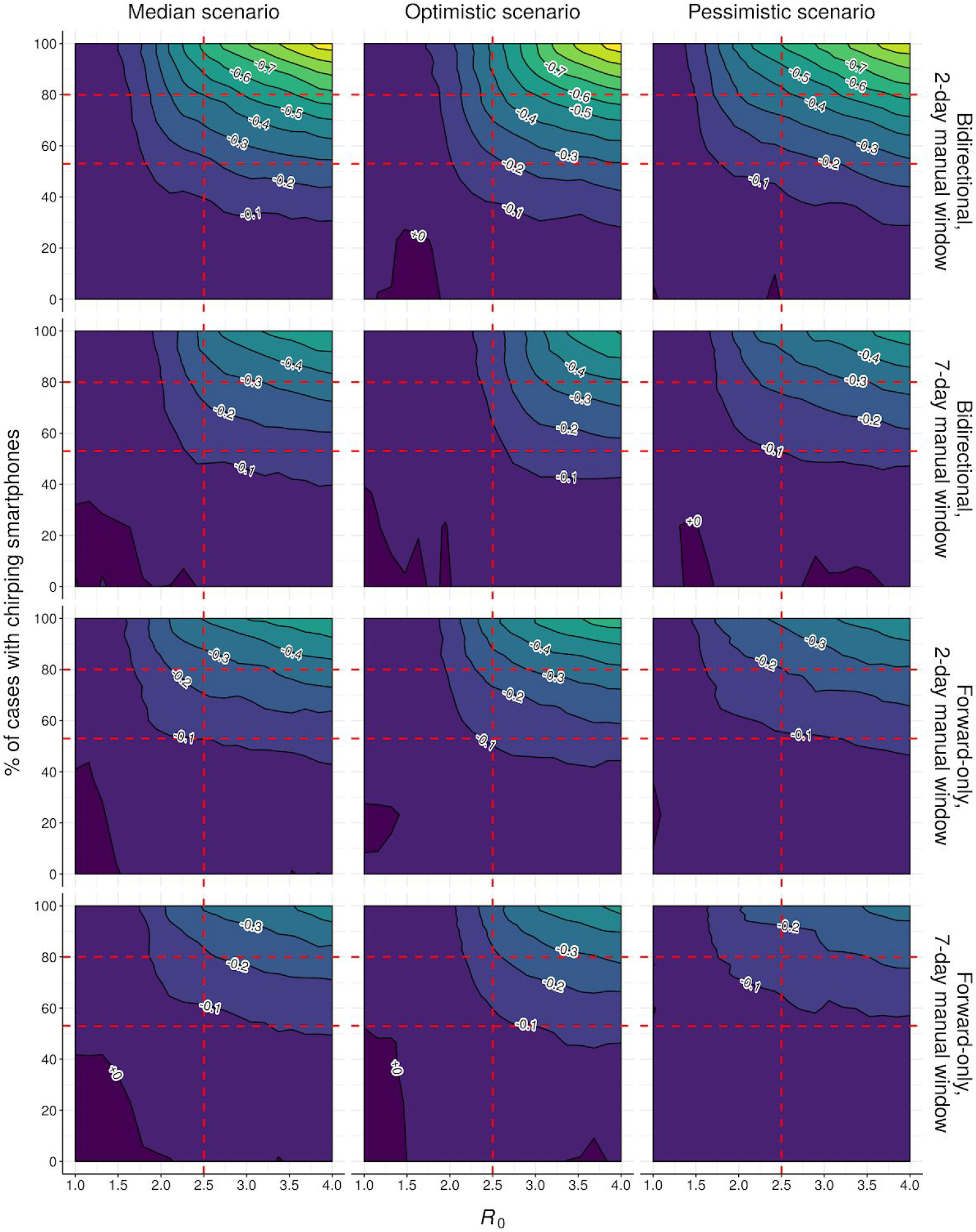
Effect of R0, network fragmentation, and disease parameters on outperformance of hybrid vs manual tracing (R_eff_). Contour plot showing difference in mean effective reproduction number (over 500 runs) between hybrid and manual-only hybrid tracing for different disease scenarios (Table S1) and tracing strategies, assuming 90% of non-environmental contacts traced and immediate tracing of symptomatic cases. Horizontal red dashed lines indicate low- and high-uptake scenarios from the main text (Table S1); vertical red dashed line indicates the *R*_0_ value used in Fig. 2 & 4. Note that the data-retention limit for the digital system in 14 days in all cases.

**Figure S23:**
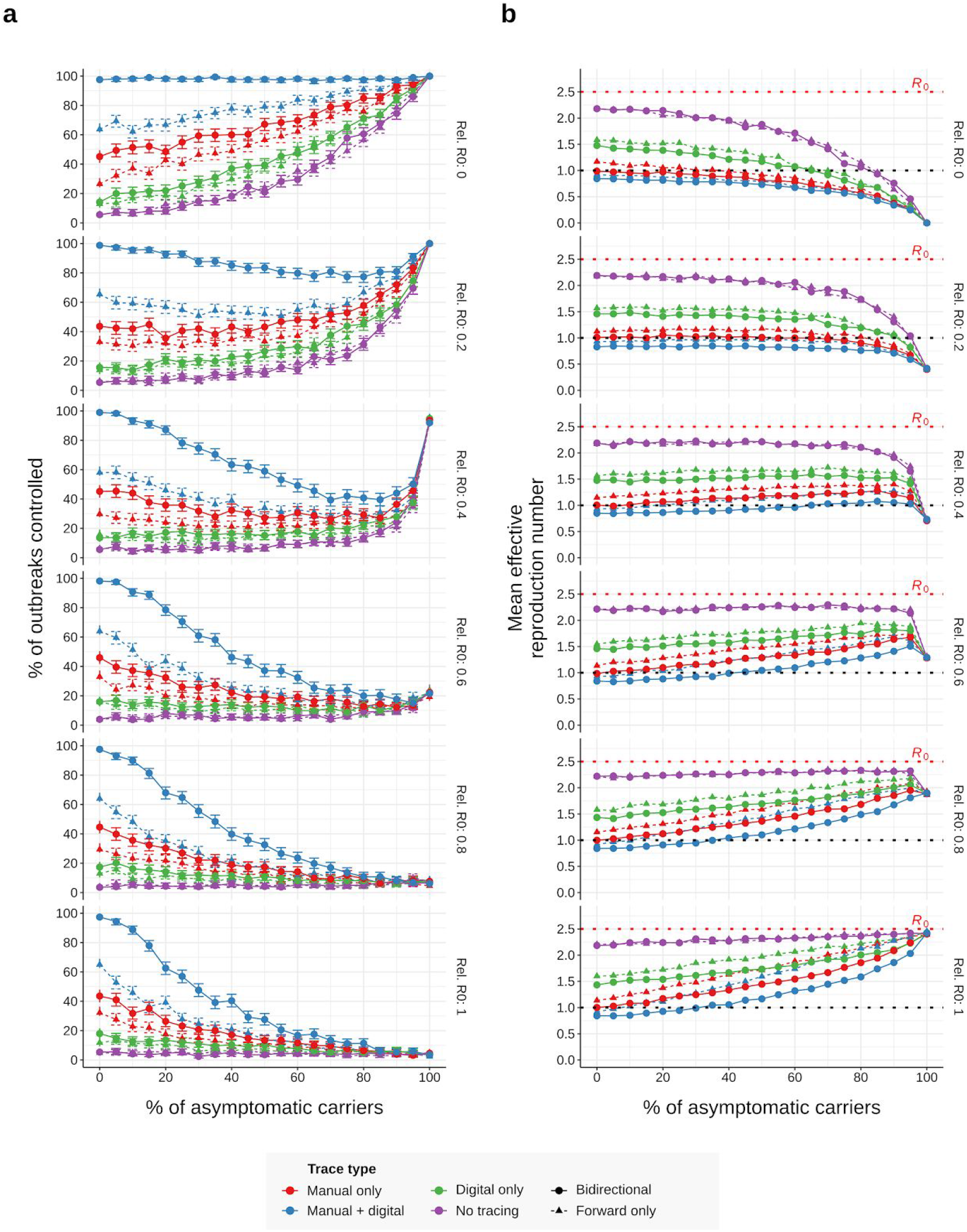
Effect of the rate and infectiousness of asymptomatic carriers on epidemic control (7-day manual window). (a) Control rates and (b) Average effective reproduction number achieved under different combinations of the rate (%) and relative infectiousness (“Rel. R0”) of asymptomatic carriers, assuming otherwise median disease parameters (Table S1), 90% of non-environmental contacts traced and a 7-day manual trace window. Error bars in (a) represent 95% credible intervals across 500 runs under a uniform beta prior; points in (b) represent average values over the same.

**Figure S24:**
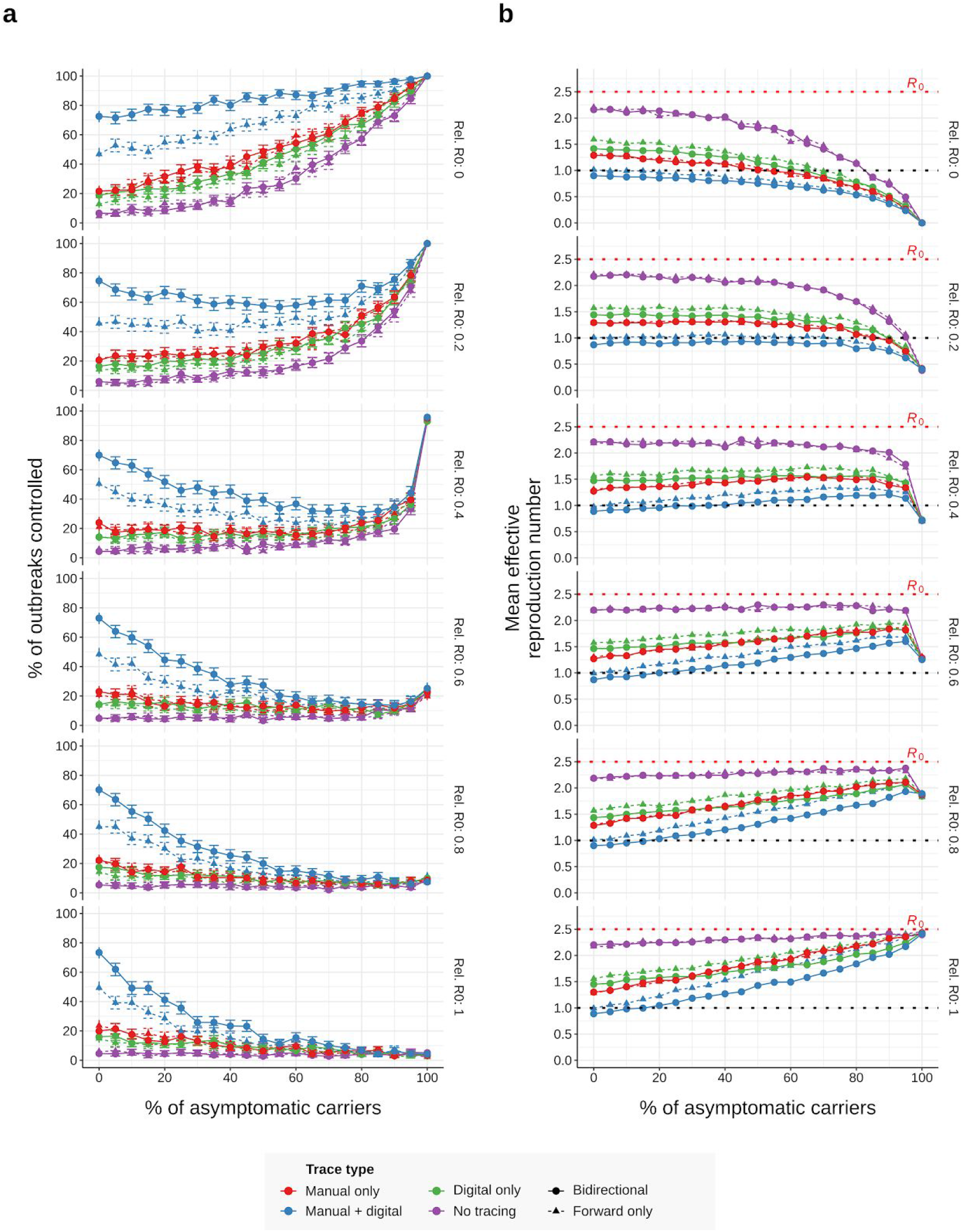
Effect of the rate and infectiousness of asymptomatic carriers on epidemic control (2-day manual window). (a) Control rates and (b) Average effective reproduction number achieved under different combinations of the rate (%) and relative infectiousness (“Rel. R0”) of asymptomatic carriers, assuming otherwise median disease parameters (Table S1), 90% of non-environmental contacts traced and a 2-day manual trace window. Error bars in (a) represent 95% credible intervals across 500 runs under a uniform beta prior; points in (b) represent average values over the same.

**Figure S25:**
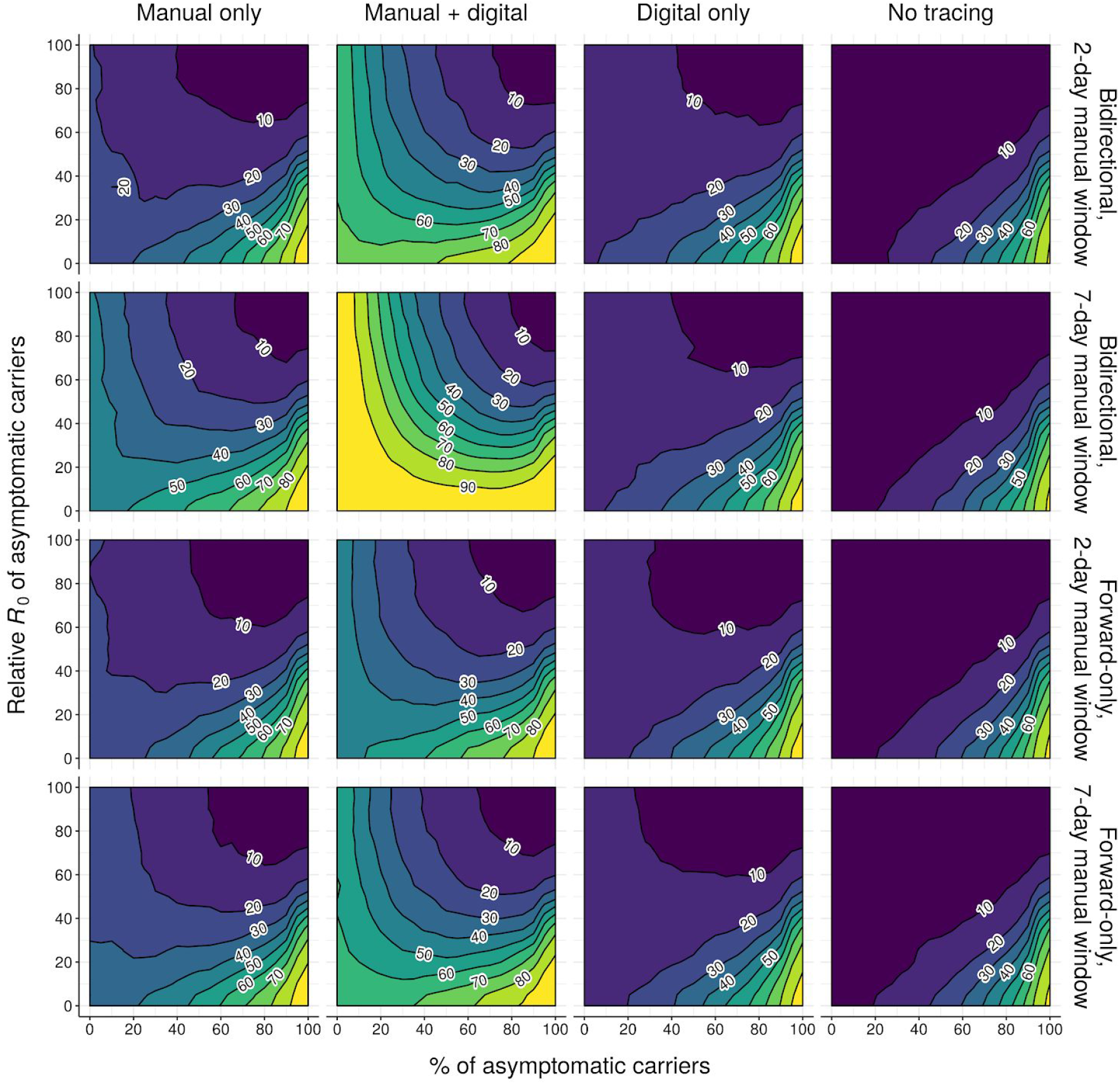
Effect of the rate and infectiousness of asymptomatic carriers on epidemic control (contour plot, control rate). Neighbour-averaged contour plots showing % of outbreaks controlled (over 500 runs) under different tracing strategies and different combinations of the rate (%) and relative infectiousness (“Relative *R*_0_”) of asymptomatic carriers, assuming otherwise median disease parameters (Table S1) and that 90% of non-environmental contacts are traced.

**Figure S26:**
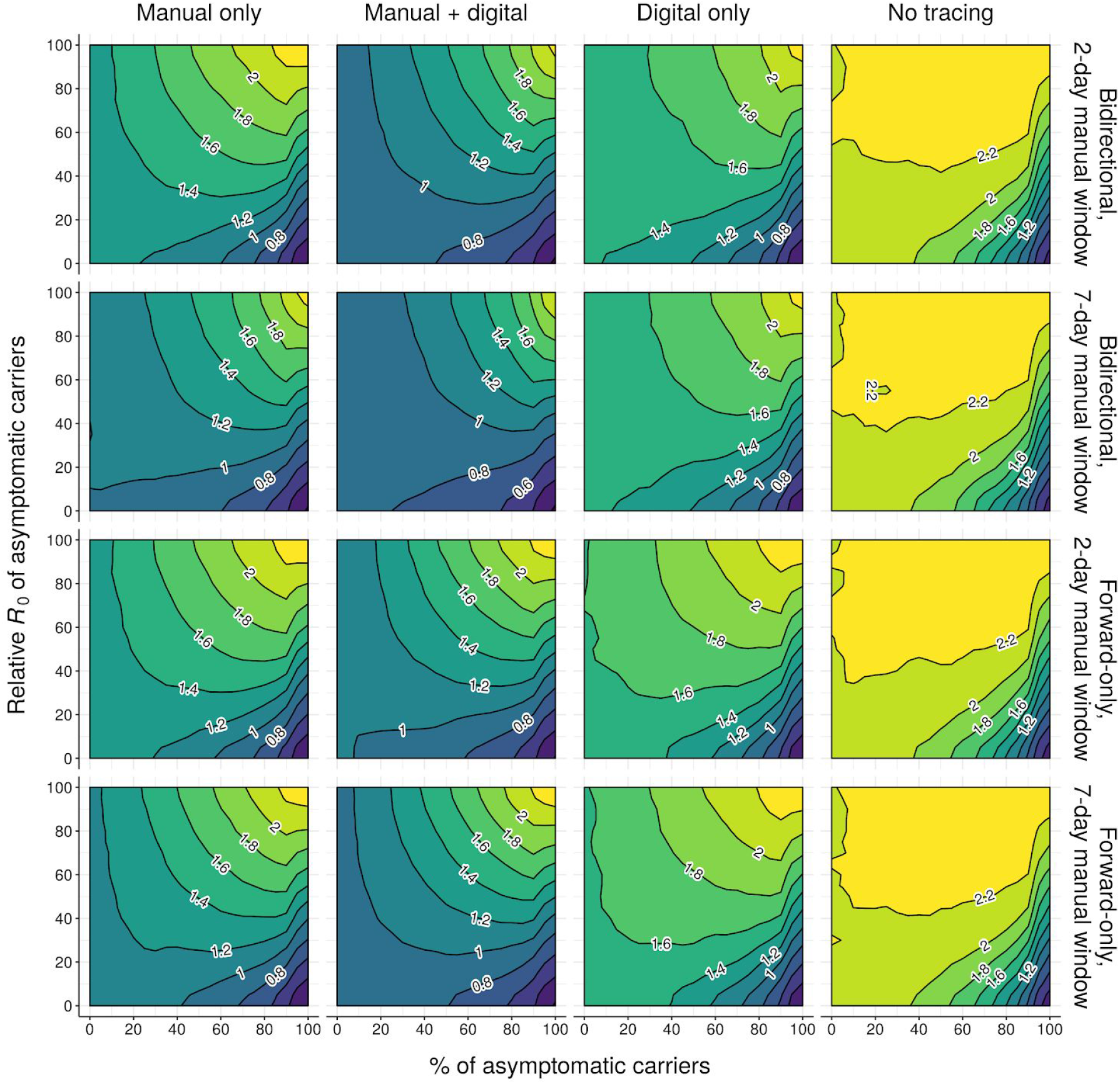
Effect of the rate and infectiousness of asymptomatic carriers on epidemic control (contour plot, *R*_eff_). Neighbour-averaged contour plots showing mean effective reproduction number (over 500 runs) achieved under different tracing strategies and different combinations of the rate (%) and relative infectiousness (“Relative *R*_0_”) of asymptomatic carriers, assuming otherwise median disease parameters (Table S1) and that 90% of non-environmental contacts are traced.

**Figure S27:**
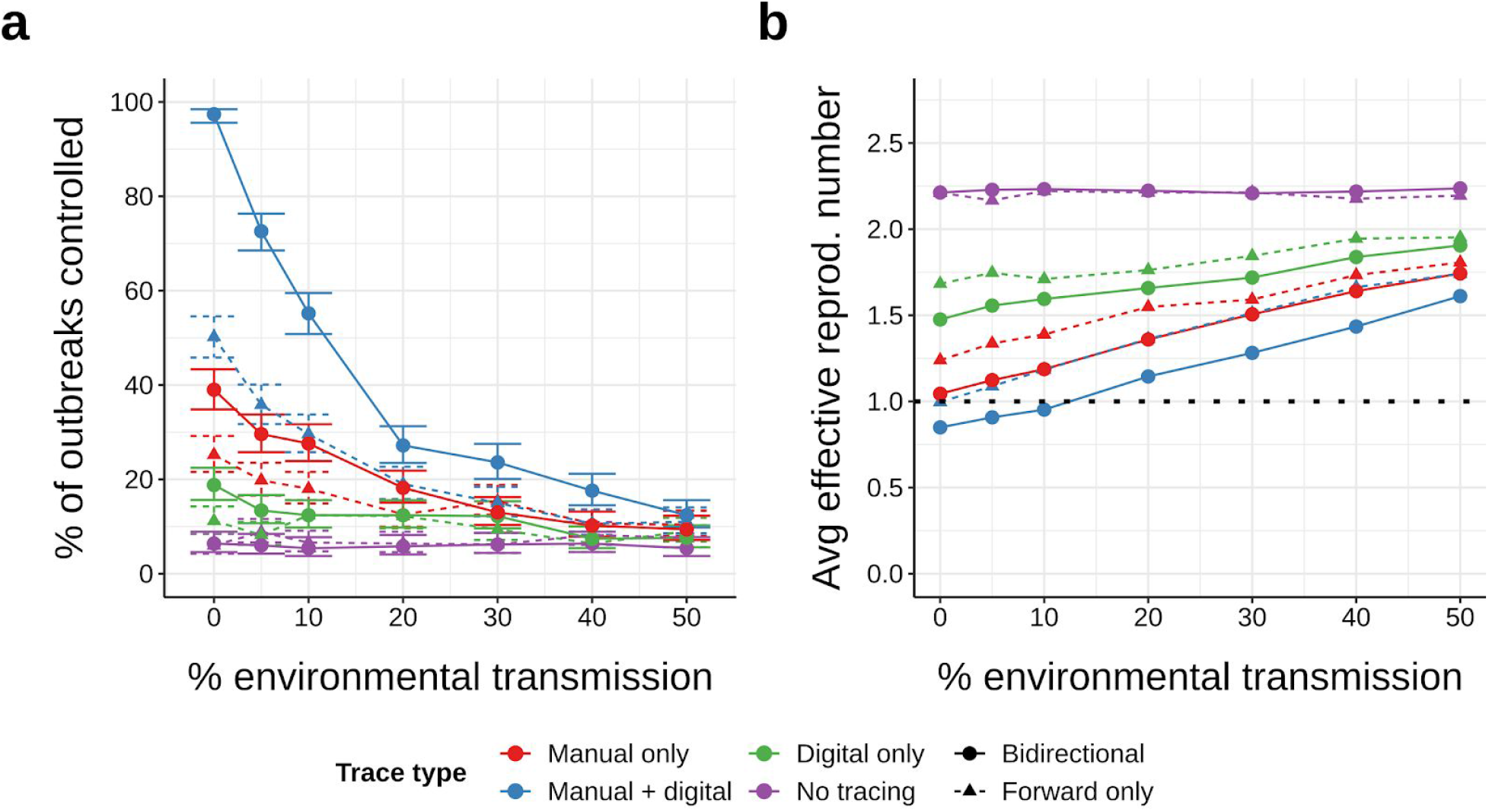
Effect of environmental transmission on epidemic control. (a) Control rates and (b) Average effective reproduction numbers achieved under different rates of environmental transmission, assuming otherwise median disease parameters (Table S1), 90% of non-environmental contacts traced and a 7-day manual trace window. Environmental transmission is assumed to be untraceable by either manual or digital contact tracing. Error bars in (a) represent 95% credible intervals across 500 runs under a uniform beta prior; points in (b) represent average values over the same.

**Figure S28:**
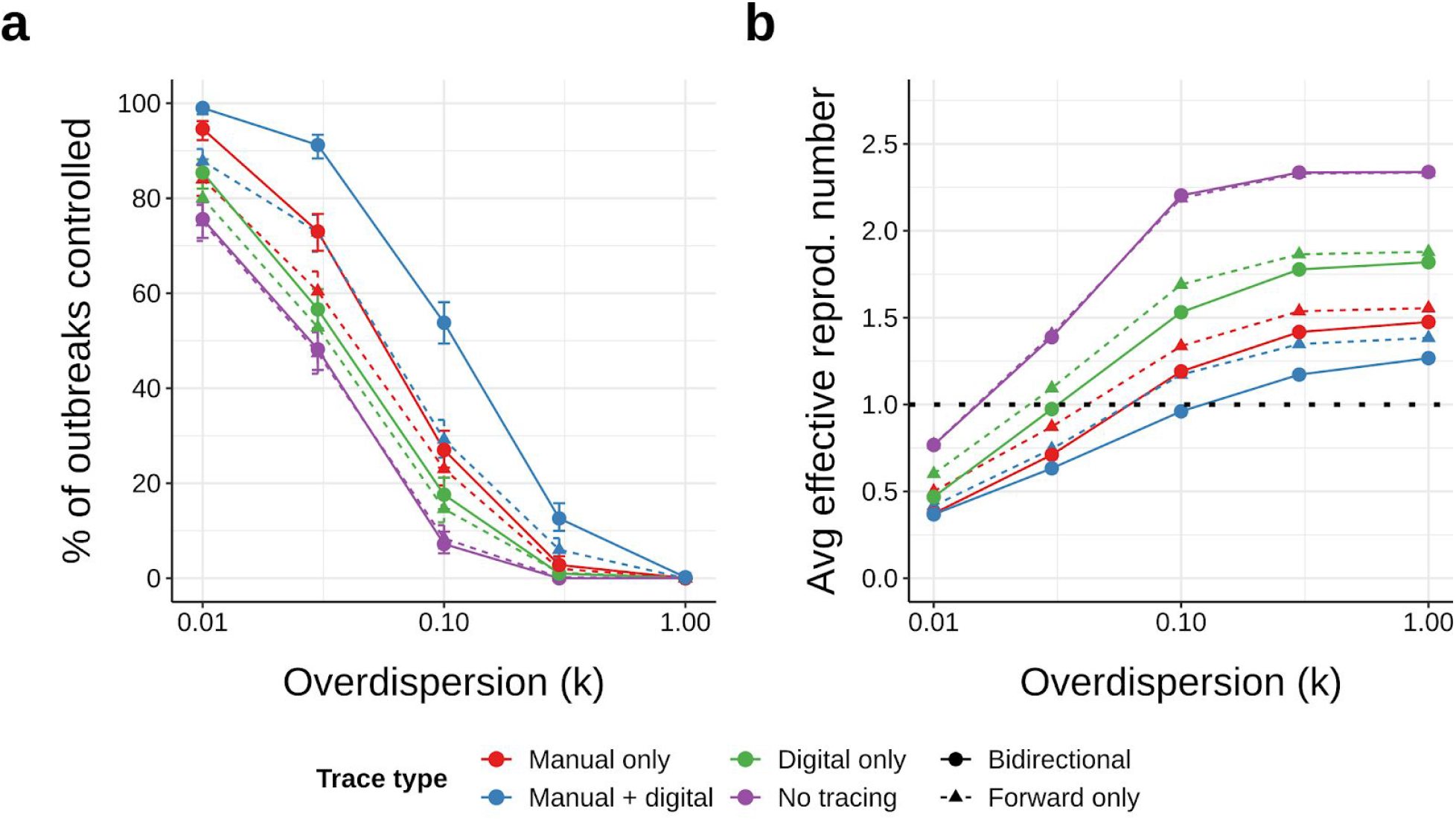
Effect of overdispersion parameter on epidemic control. (a) Control rates and (b) Average effective reproduction numbers achieved under different values of the overdispersion parameter *k* (where lower values of *k* denote higher variance in infectiousness among cases), assuming otherwise median disease parameters (Table S1), 90% of non-environmental contacts traced and a 7-day manual trace window. Error bars in (a) represent 95% credible intervals across 500 runs under a uniform beta prior; points in (b) represent average values over the same.

**Figure S29:**
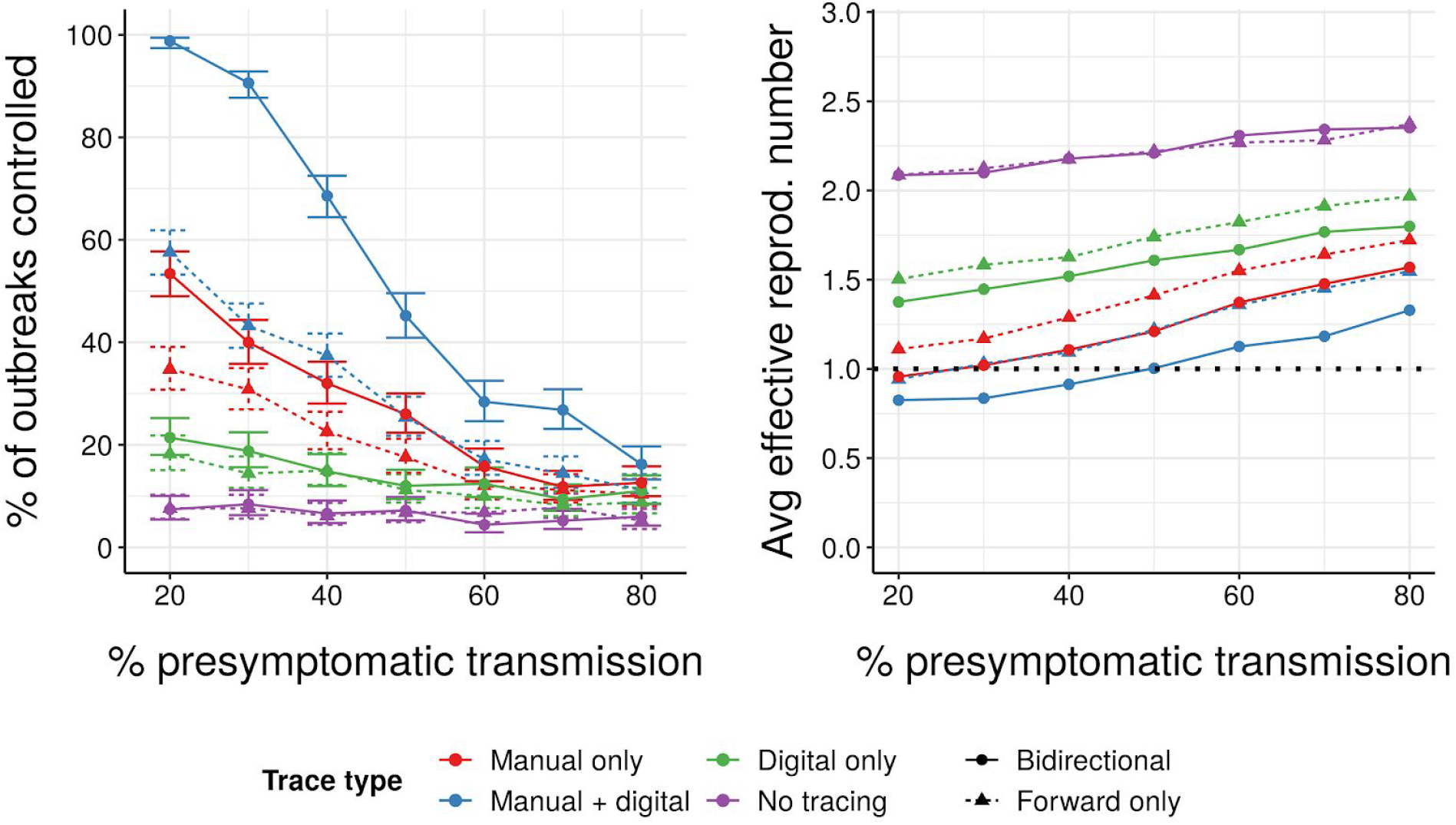
Effect of pre-symptomatic transmission on epidemic control. (a) Control rates and (b) Average effective reproduction numbers achieved under different rates of pre-symptomatic transmission, assuming otherwise median disease parameters (Table S1), 90% of non-environmental contacts traced and a 7-day manual trace window. Error bars in (a) represent 95% credible intervals across 500 runs under a uniform beta prior; points in (b) represent average values over the same.

**Figure S30:**
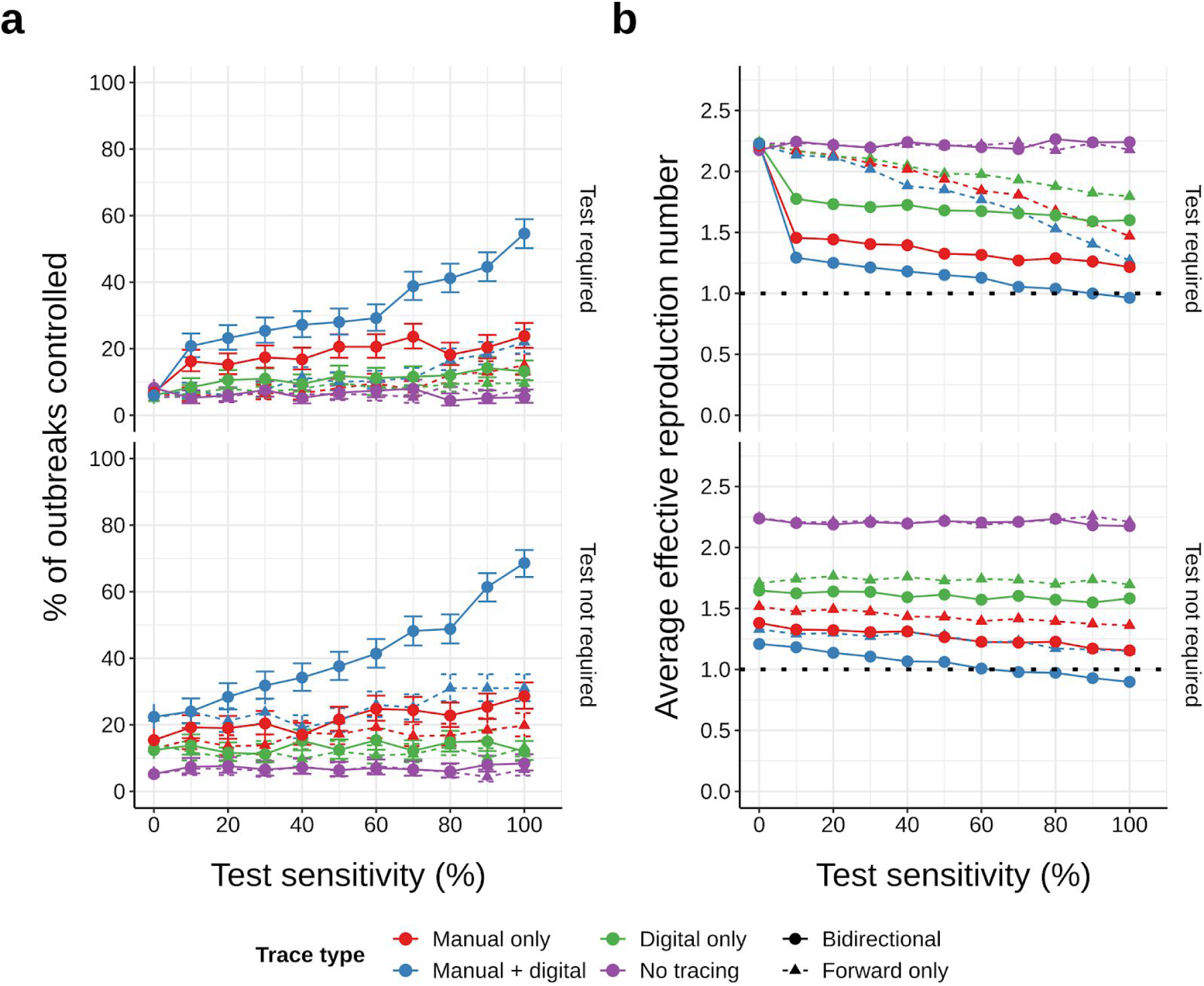
Effect of test sensitivity on epidemic control. (a) Control rates and (b) Average effective reproduction numbers achieved under different assumptions about test sensitivity (x-axis) and whether or not a positive test result is required before contact tracing from a symptomatic case (top/bottom panels), assuming otherwise median disease parameters (Table S1), 90% of non-environmental contacts traced and a 7-day manual trace window. Error bars in (a) represent 95% credible intervals across 500 runs under a uniform beta prior; points in (b) represent average values over the same.

**Figure S31:**
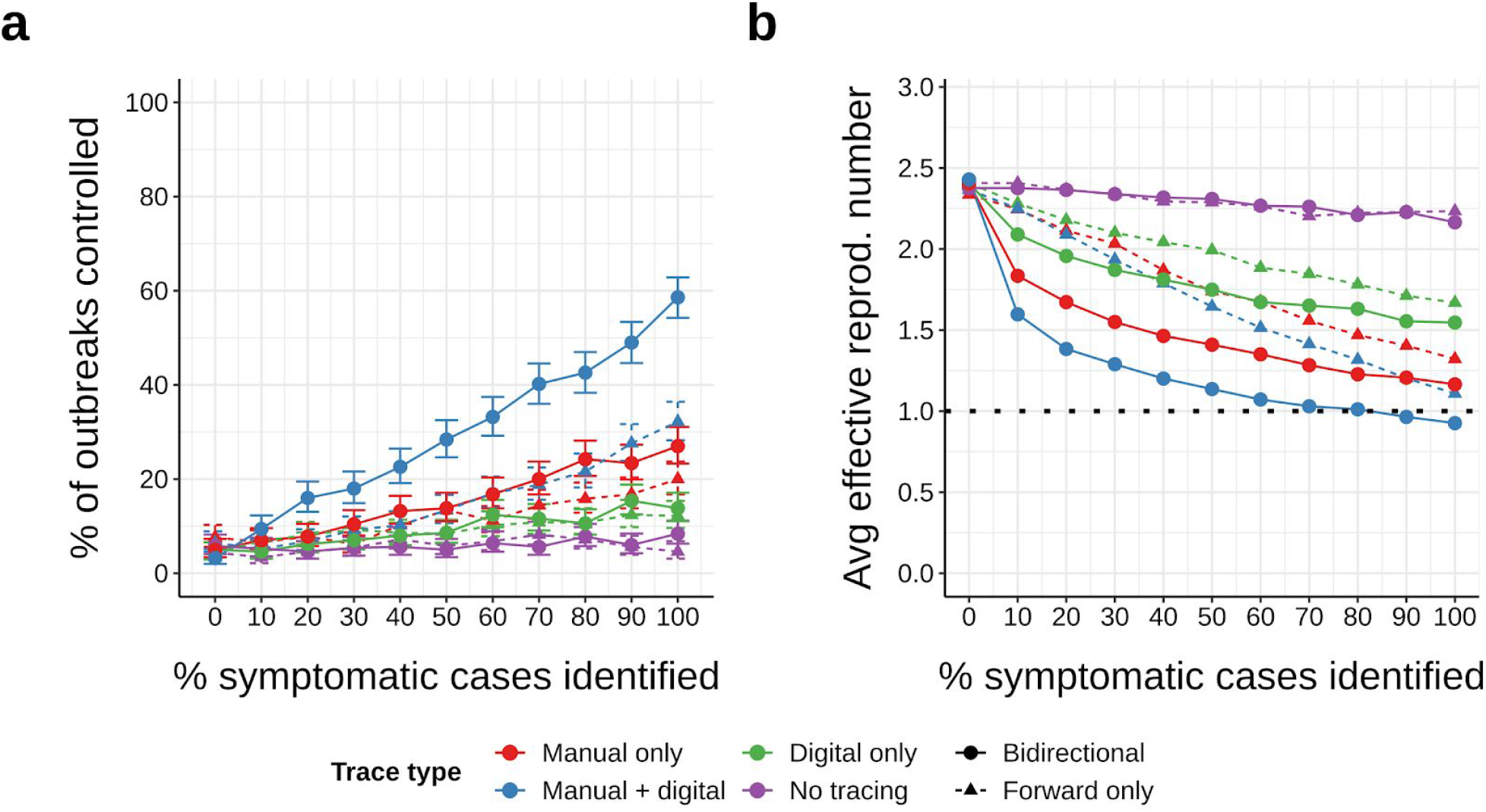
Effect of symptomatic case ascertainment on epidemic control. (a) Control rates and (b) average effective reproduction numbers achieved as a function of the probability of identifying symptomatic cases (based on symptoms alone), assuming otherwise median disease parameters (Table S1), 90% probability of trace success, and a 7-day manual trace window. Error bars in (a) represent 95% credible intervals across 500 runs under a uniform beta prior; points in (b) represent average values over the same. Note that, since 45% of cases are asymptomatic (and thus never identified from symptoms), overall ascertainment when *x*% of symptomatic cases are identified is roughly 0.55*x*%.

**Figure S32:**
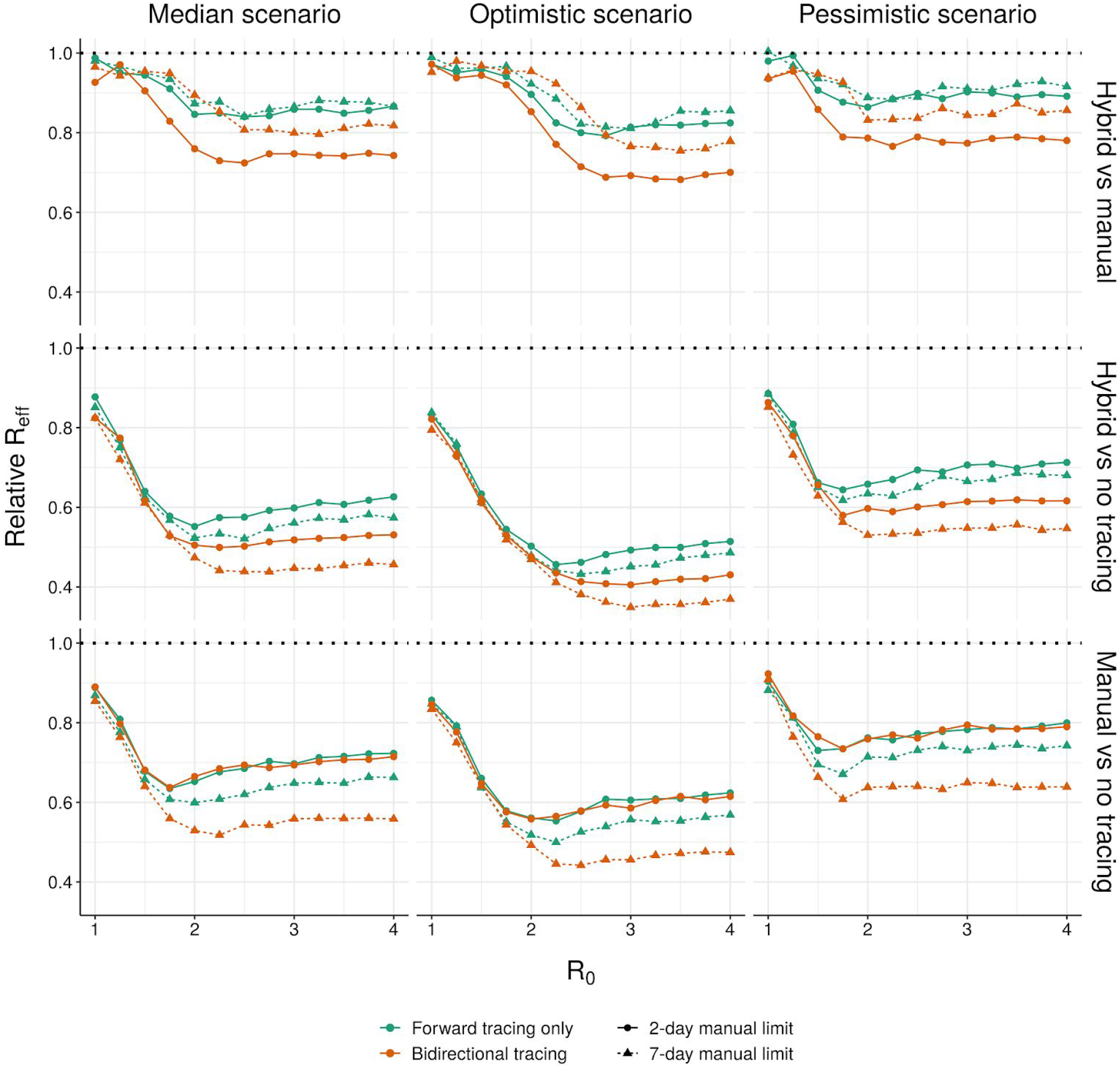
Relative effective reproduction numbers of hybrid, manual and no tracing. Ratio between the effective reproduction number (over 1000 runs per condition) achieved by (top row) hybrid and manual tracing, (middle row) hybrid and no tracing, (bottom row) manual and no tracing, under (left) median, (middle) optimistic and (right) pessimistic disease parameters (Table S1), given 90% of non-environmental contacts traced, high uptake of the digital system, and immediate tracing of symptomatic cases.

**Figure S33.**
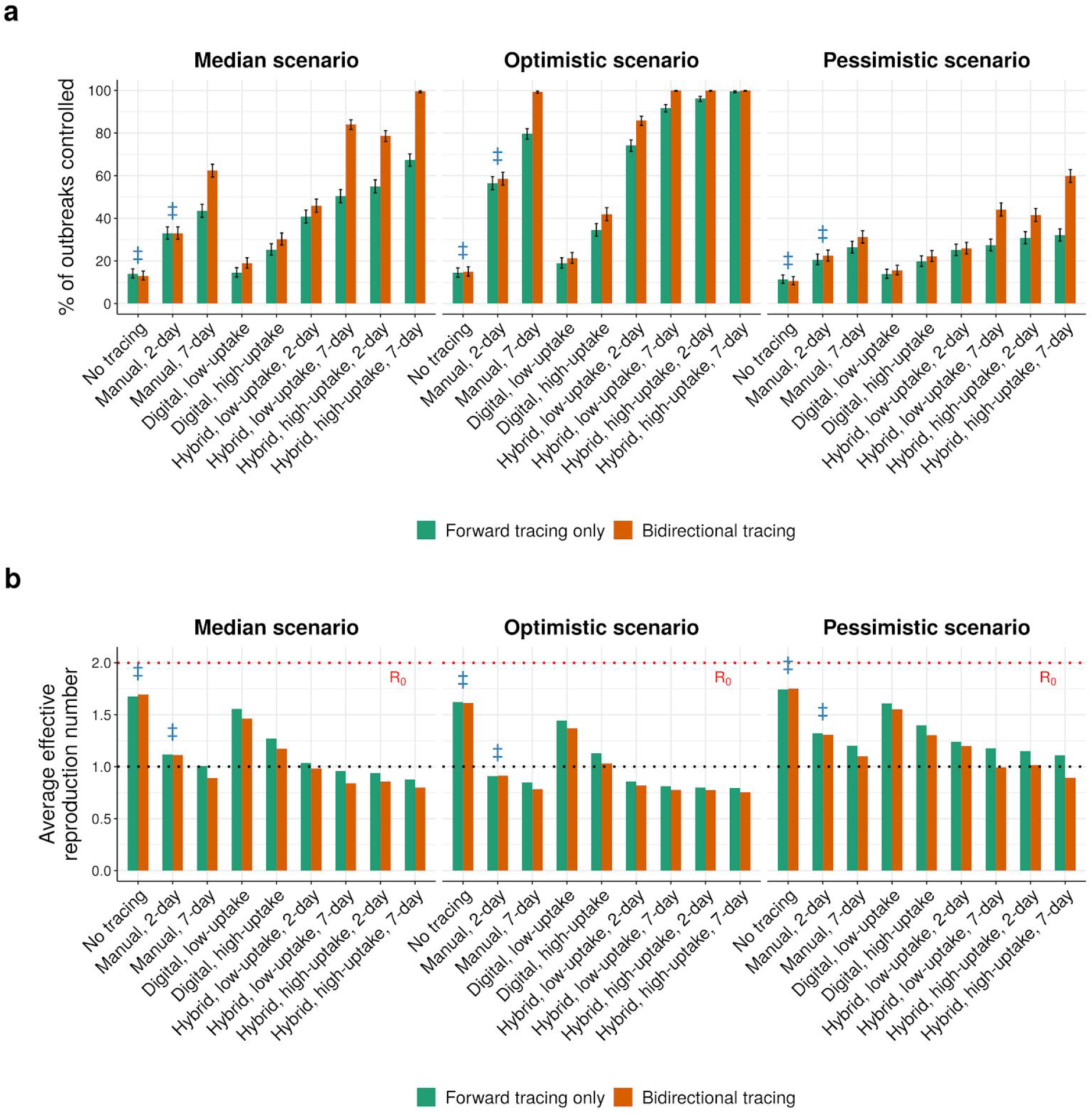
Tracing strategies for controlling COVID-19 (*R*_0_ = 2.0). As Fig. 4, but assuming an *R*0 of 2.0. Blue ‡ symbols indicate current practice in most regions. Low and high uptake correspond to 53% and 80% of cases, respectively, having chirp-enabled smartphones. Error bars in (a) represent 95% credible intervals across 1000 runs under a uniform beta prior. Without tracing, forward and bidirectional are equivalent.

**Figure S34.**
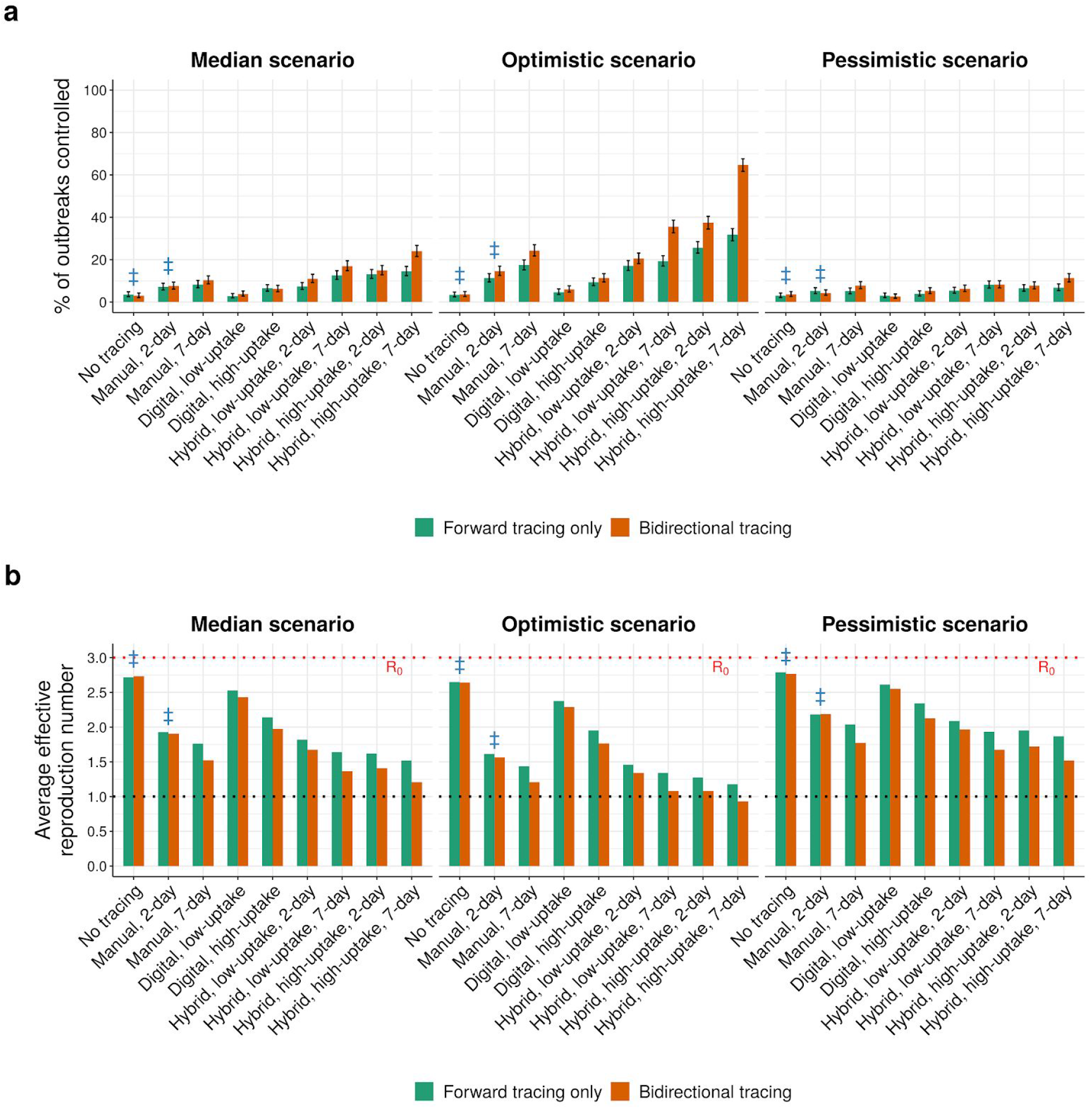
Tracing strategies for controlling COVID-19 (*R*_0_ = 3.0). As Fig. 4, but assuming an *R*0 of 3.0. Blue ‡ symbols indicate current practice in most regions. Low and high uptake correspond to 53% and 80% of cases, respectively, having chirp-enabled smartphones. Error bars in (a) represent 95% credible intervals across 1000 runs under a uniform beta prior. Without tracing, forward and bidirectional are equivalent.

**Figure S35.**
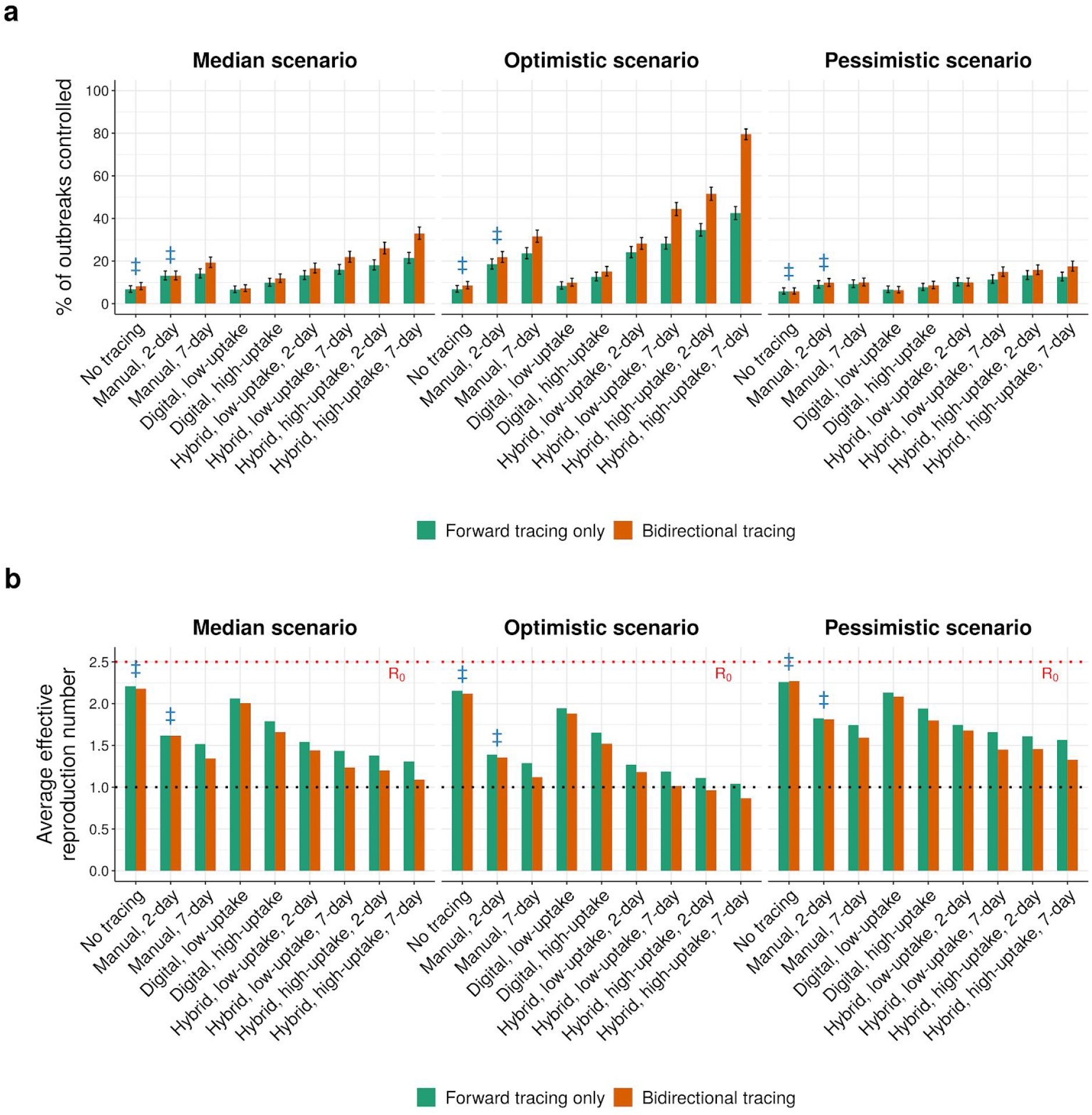
Tracing strategies for controlling COVID-19 (80% contacts traced). As Fig. 4, but assuming only 80% of non-environmental contacts are traced. Blue ‡ symbols indicate current practice in most regions. Low and high uptake correspond to 53% and 80% of cases, respectively, having chirp-enabled smartphones. Error bars in (a) represent 95% credible intervals across 1000 runs under a uniform beta prior. Without tracing, forward and bidirectional are equivalent.

**Figure S36.**
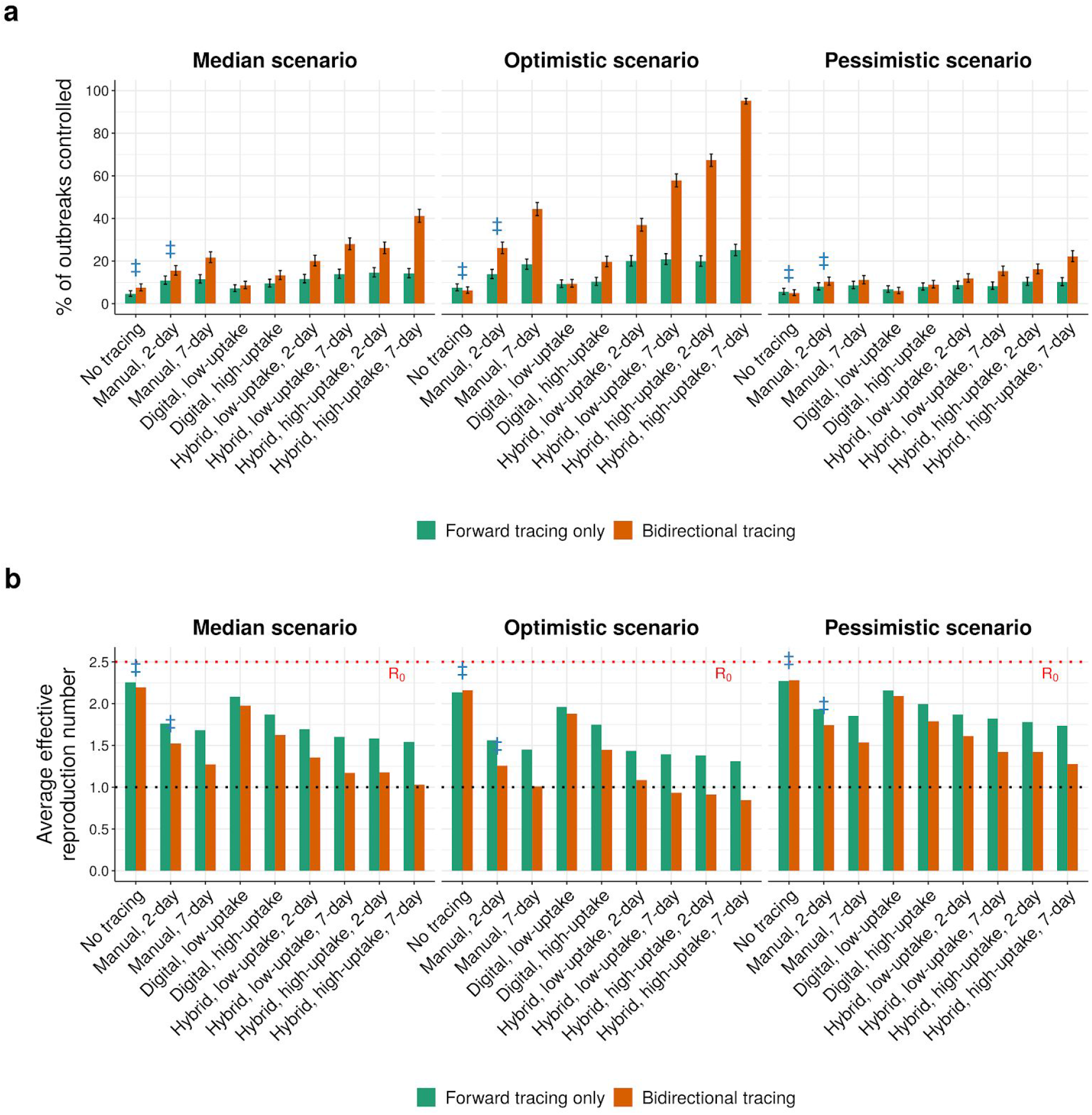
Tracing strategies for controlling COVID-19 (pre-emptive testing). As Fig. 4, but assuming that symptomatic cases require a positive test before their contacts are traced. Blue ‡ symbols indicate current practice in most regions. Low and high uptake correspond to 53% and 80% of cases, respectively, having chirp-enabled smartphones. Error bars in (a) represent 95% credible intervals across 1000 runs under a uniform beta prior. Without tracing, forward and bidirectional are equivalent.

